# The impact of complement factor H-related protein gene deletions on kidney transplantation

**DOI:** 10.1101/2024.02.18.24301068

**Authors:** Markkinen Salla, Lokki A. Inkeri, Helanterä Ilkka, Ritari Jarmo, Partanen Jukka, Meri Seppo, Hyvärinen Kati

## Abstract

We recently reported that a homozygous deletion in the complement factor H-related (*CFHR)* locus predisposed kidney transplant patients to rejection. As donors carried intact genes, the susceptibility may have resulted from alloimmune reaction to FHR proteins. However, we found no evidence for anti-FH response. It is therefore possible that *CFHR* deletions as such affect the rejection risk. Here, we used MLPA and WGS to fine-map and sequence the *CFHR* region in rs7542235-GG patients, a SNP tagging for ΔCFHR311 deletion. Our results confirmed that all patients with this SNP harboured deletions of various sizes encompassing *CFHR1*. Furthermore, patients with homozygous ΔCFHR311 were homozygous for rs6677604-A, a SNP tagging for deletions of CFHR311 locus, confirming that allele A tags for deletion of both *CFHR3* and *CFHR1*. Proteomics analyses in a larger population demonstrated that rs7542235-G and rs6677604-A associate with expression levels of several proteins involved in regulating alloimmune response. We observed that while increasing the rejection risk, the ΔCFHR311 did not associate to baseline disease or specific clinical characteristics. To conclude, the various deletion types found in patients shared the deletion of *CFHR1* gene confirming its association to variant rs7542235. Also, both deletion-tagging alleles are associated with altered expression of FHR proteins.

## INTRODUCTION

Prior matching of human leukocyte antigen (HLA), ABO blood group types, and a negative anti-HLA cross-matching are the golden rule in kidney transplantation. Regardless, some of the patients still experience complications after kidney transplantation. In search of novel histocompatibility factors, a study by Steers *et al*.(1) reported that a homozygous gene deletion of *LIMS1*, and our previous study (2) reported that homozygous deletion in the complement factor H-related (*CFHR)* locus consisting of *CFHR3* and *CFHR1* genes were associated with acute rejection of the graft.

There are a few mutually non-exclusive mechanistic explanations to these findings. The rejection may be related to alloimmune response by the patient against the protein missing from the patient but expressed by the graft, hence, as such not necessarily linked to the function of the protein. Alternatively, the rejection risk may be related to the function of the protein missing from the patient. The functional effect can be direct, i.e., a result of the missing protein, or caused by expression of the missing protein by the transplanted kidney, or it can be related to complex downstream networks. A deletion of a gene and adjacent DNA segments may furthermore lead to changes in the expression of some other genes that can be identified, for example, using quantitative trait loci (QTL) (3–5) approaches.

Steers and co-workers, who reported association between *LIMS1* gene deletion and graft rejection in kidney transplantation, could identify a specific anti-LIMS1 antibody pointing to alloimmune response against the LIMS1 protein missing from the patient. In our recent study (6), we could not replicate the *LIMS1* deletion association in the set of 1025 Finnish kidney transplantation patients and donors, but instead reported an association between *CFH/CFHR* locus deletion and acute rejection. The *CFH/CFHR* locus contains genes encoding for factor H (FH), a major regulator of the alternative pathway (AP) of complement activation, and a set of related genes, *CFHR1–5* (6,7). FH inhibits AP activation both in body fluids and on cell surfaces. It binds to C3b, which has become bound to self surfaces, where it inhibits local complement activation. In contrast, on “foreign” targets, such as microbial or virus surfaces, binding of FH to surface deposited C3b is weak allowing AP activation and amplification to occur (8–10). The functions of FHR-1–5 proteins are becoming better understood. As suggested already a long time ago (11) the FHRs promote complement activity by competing with the master inhibitor FH (7). Genetic variants of *CFH* and *CFHR1–5* have been linked to the pathogenesis of some kidney diseases, such as C3 glomerulopathy (12), atypical hemolytic uremic syndrome (13,14), IgA nephropathy (15–17) and lupus nephritis (18).

In the present study, we wanted to understand the effects of Δ*CFHR3–1* gene deletions in detail. We fine-mapped the deletion boundaries using multiple ligation-dependent probe amplification (MLPA) and whole genome sequencing (WGS). We found that deletions of various sizes shared a homozygous deletion of *CFHR1* gene indicating that rs7542235 allele G tags for a heterozygous deletion of that locus. In addition to rs7542235, previous studies have shown that another variant, rs6677604, tags for a deletion at the same genomic region (16–18). We observed that a rs6677604 allele A tags for a heterozygous deletion of *CFHR3–1* locus. Proteome analysis of patient plasma samples revealed that, in fact, the deletion-tagging alleles rs7542235-G and rs6677604-A correlated with plasma protein level changes of at least 23 and seven proteins, respectively, sharing six proteins. These shortlists merit further studies.

## MATERIALS AND METHODS

### Study cohort

The present study included 15 patients from the study of Markkinen *et al*. (6), who had rs7542235 GG-genotype tagging for the homozygous deletion in the Δ*CFHR3–1* locus. The patients received their first kidney transplantation during 2007–2017 at the Helsinki University Hospital, Helsinki, Finland. The characteristics of the whole study population including the 15 patients with homozygous rs7542235-G and 1010 patients with heterozygous (AG) or homozygous non-deletion genotype (AA) are presented in **Table 1**. In total of 199 (20%) rejections were observed in our study cohort. The primary outcome for these patients was biopsy-proven acute rejection, including both antibody-mediated and T-cell-mediated rejections (19). Borderline changes were also included as rejections. The characteristics of the 15 patients (8 in the rejection-group and 7 in the non-rejection-group) with Δ*CFHR3–1*-deletion tagging rs7542235 GG-genotype are presented in **Table 2**.

**Table 1.**
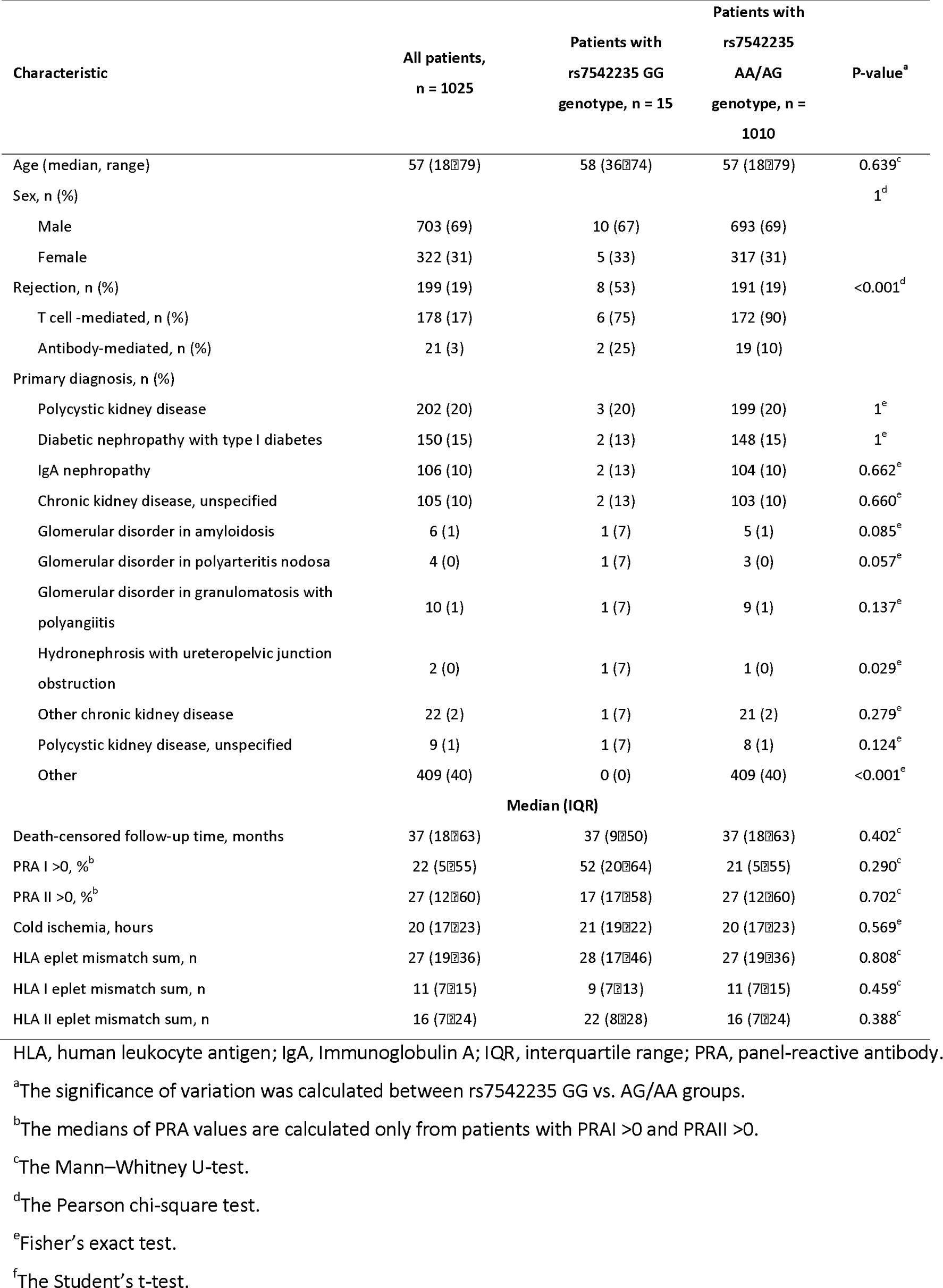
Characteristics of the study cohort.

| Characteristic | All patients,<br>n = 1025 | Patients with<br>rs7542235 GG<br>genotype, n = 15 | Patients with<br>rs7542235<br>AA/AG<br>genotype, n =<br>1010 | P-value<sup>a</sup> |
| --- | --- | --- | --- | --- |
| Age (median, range) | 57 (18–79) | 58 (36–74) | 57 (18–79) | 0.639<sup>c</sup> |
| Sex, n (%) |  |  |  | 1<sup>d</sup> |
| Male | 703 (69) | 10 (67) | 693 (69) |  |
| Female | 322 (31) | 5 (33) | 317 (31) |  |
| Rejection, n (%) | 199 (19) | 8 (53) | 191 (19) | <0.001<sup>d</sup> |
| T cell -mediated, n (%) | 178 (17) | 6 (75) | 172 (90) |  |
| Antibody-mediated, n (%) | 21 (3) | 2 (25) | 19 (10) |  |
| Primary diagnosis, n (%) |  |  |  |  |
| Polycystic kidney disease | 202 (20) | 3 (20) | 199 (20) | 1<sup>e</sup> |
| Diabetic nephropathy with type1 diabetes | 150 (15) | 2 (13) | 148 (15) | 1<sup>e</sup> |
| IgA nephropathy | 106 (10) | 2 (13) | 104 (10) | 0.662<sup>e</sup> |
| Chronic kidney disease, unspecified | 105 (10) | 2 (13) | 103 (10) | 0.660<sup>e</sup> |
| Glomerular disorder in amyloidosis | 6 (1) | 1 (7) | 5 (1) | 0.085<sup>e</sup> |
| Glomerular disorder in polyarteritis nodosa | 4 (0) | 1 (7) | 3 (0) | 0.057<sup>e</sup> |
| Glomerular disorder in granulomatosis with<br>polyangiitis | 10 (1) | 1 (7) | 9 (1) | 0.137<sup>e</sup> |
| Hydronephrosis with ureteropelvic junction<br>obstruction | 2 (0) | 1 (7) | 1 (0) | 0.029<sup>e</sup> |
| Other chronic kidney disease | 22 (2) | 1 (7) | 21 (2) | 0.279<sup>e</sup> |
| Polycystic kidney disease, unspecified | 9 (1) | 1 (7) | 8 (1) | 0.124<sup>e</sup> |
| Other | 409 (40) | 0 (0) | 409 (40) | <0.001<sup>e</sup> |
| **Median (IQR)** |  |  |  |  |
| Death-censored follow-up time, months | 37 (18–63) | 37 (9–50) | 37 (18–63) | 0.402<sup>c</sup> |
| PRA I >0, %<sup>b</sup> | 22 (5–55) | 52 (20–64) | 21 (5–55) | 0.290<sup>c</sup> |
| PRA II >0, %<sup>b</sup> | 27 (12–60) | 17 (17–58) | 27 (12–60) | 0.702<sup>c</sup> |
| Cold ischemia, hours | 20 (17–23) | 21 (19–22) | 20 (17–23) | 0.569<sup>e</sup> |
| HLA eplet mismatch sum, n | 27 (19–36) | 28 (17–46) | 27 (19–36) | 0.808<sup>c</sup> |
| HLA I eplet mismatch sum, n | 11 (7–15) | 9 (7–13) | 11 (7–15) | 0.459<sup>c</sup> |
| HLA II eplet mismatch sum, n | 16 (7–24) | 22 (8–28) | 16 (7–24) | 0.388<sup>c</sup> |

HLA, human leukocyte antigen; IgA, Immunoglobulin A; IQR, interquartile range; PRA, panel-reactive antibody.
<sup>a</sup>The significance of variation was calculated between rs7542235 GG vs. AG/AA groups.
<sup>b</sup>The medians of PRA values are calculated only from patients with PRA I >0 and PRA II >0.
<sup>c</sup>The Mann–Whitney U-test.
<sup>d</sup>The Pearson chi-square test.
<sup>e</sup>Fisher's exact test.
<sup>f</sup>The Student's t-test.

**Table 2.**
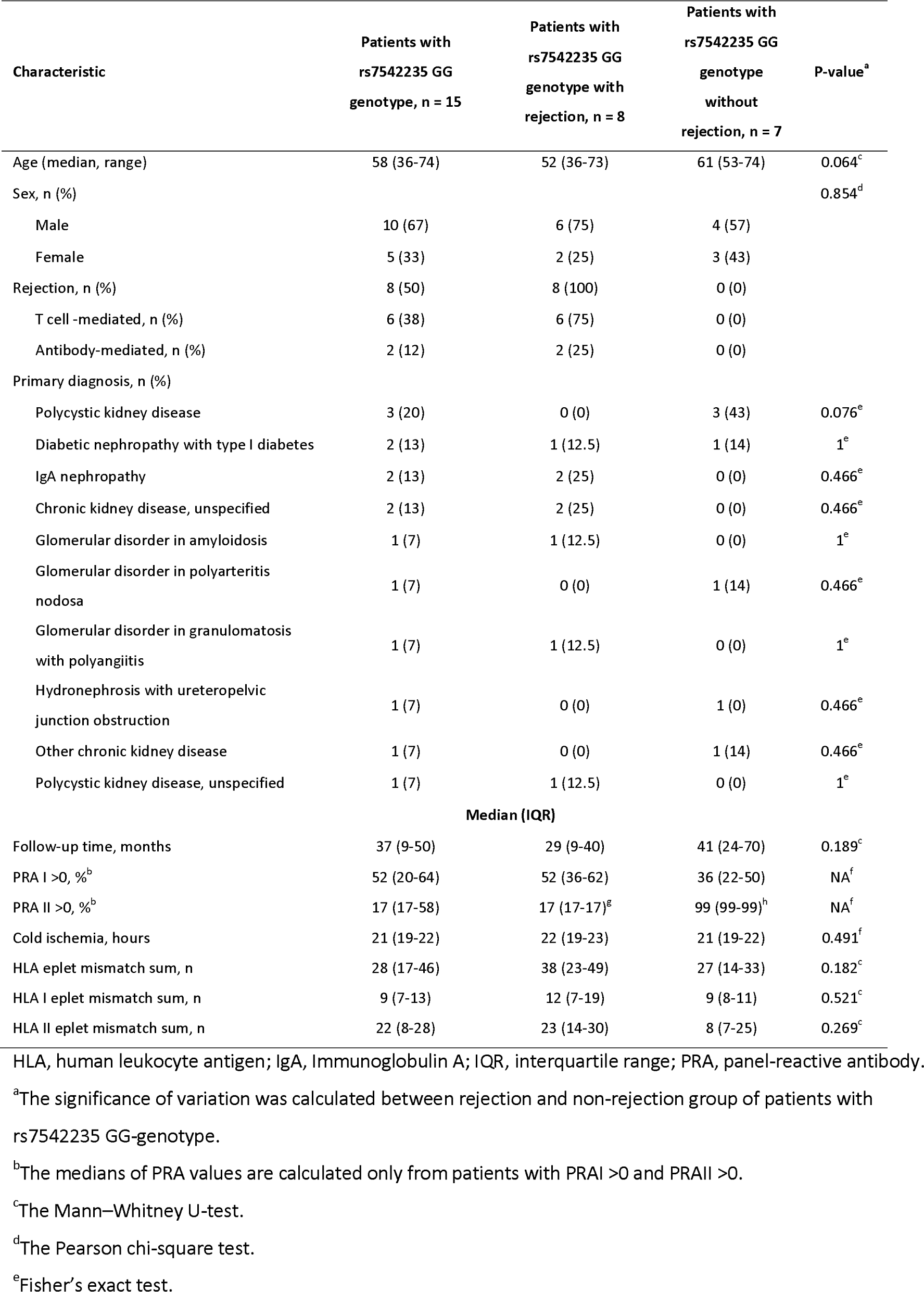
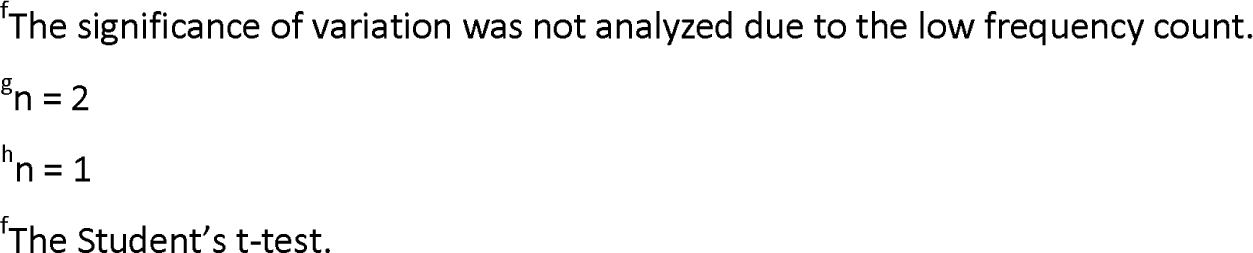
Characteristics of the 15 patients with rs7542235 GG-genotype tagging for homozygous *CFHR*3⍰1 deletion. Rejection vs. non-rejection group.

| Characteristic | Patients with rs7542235 GG genotype, n = 15 | Patients with rs7542235 GG genotype with rejection, n = 8 | Patients with rs7542235 GG genotype without rejection, n = 7 | P-value<sup>a</sup> |
| --- | --- | --- | --- | --- |
| Age (median, range) | 58 (36-74) | 52 (36-73) | 61 (53-74) | 0.064<sup>c</sup> |
| Sex, n (%) |  |  |  | 0.854<sup>d</sup> |
| Male | 10 (67) | 6 (75) | 4 (57) |  |
| Female | 5 (33) | 2 (25) | 3 (43) |  |
| Rejection, n (%) | 8 (50) | 8 (100) | 0 (0) |  |
| T cell-mediated, n (%) | 6 (38) | 6 (75) | 0 (0) |  |
| Antibody-mediated, n (%) | 2 (12) | 2 (25) | 0 (0) |  |
| Primary diagnosis, n (%) |  |  |  |  |
| Polycystic kidney disease | 3 (20) | 0 (0) | 3 (43) | 0.076<sup>e</sup> |
| Diabetic nephropathy with type I diabetes | 2 (13) | 1 (12.5) | 1 (14) | 1<sup>e</sup> |
| IgA nephropathy | 2 (13) | 2 (25) | 0 (0) | 0.466<sup>e</sup> |
| Chronic kidney disease, unspecified | 2 (13) | 2 (25) | 0 (0) | 0.466<sup>e</sup> |
| Glomerular disorder in amyloidosis | 1 (7) | 1 (12.5) | 0 (0) | 1<sup>e</sup> |
| Glomerular disorder in polyarteritis nodosa | 1 (7) | 0 (0) | 1 (14) | 0.466<sup>e</sup> |
| Glomerular disorder in granulomatosis with polyangiitis | 1 (7) | 1 (12.5) | 0 (0) | 1<sup>e</sup> |
| Hydronephrosis with ureteropelvic junction obstruction | 1 (7) | 0 (0) | 1 (0) | 0.466<sup>e</sup> |
| Other chronic kidney disease | 1 (7) | 0 (0) | 1 (14) | 0.466<sup>e</sup> |
| Polycystic kidney disease, unspecified | 1 (7) | 1 (12.5) | 0 (0) | 1<sup>e</sup> |
| Median (IQR) |  |  |  |  |
| Follow-up time, months | 37 (9-50) | 29 (9-40) | 41 (24-70) | 0.189<sup>c</sup> |
| PRA I >0, %<sup>b</sup> | 52 (20-64) | 52 (36-62) | 36 (22-50) | NA<sup>f</sup> |
| PRA II >0, %<sup>b</sup> | 17 (17-58) | 17 (17-17)<sup>g</sup> | 99 (99-99)<sup>h</sup> | NA<sup>f</sup> |
| Cold ischemia, hours | 21 (19-22) | 22 (19-23) | 21 (19-22) | 0.491<sup>f</sup> |
| HLA eplet mismatch sum, n | 28 (17-46) | 38 (23-49) | 27 (14-33) | 0.182<sup>c</sup> |
| HLA I eplet mismatch sum, n | 9 (7-13) | 12 (7-19) | 9 (8-11) | 0.521<sup>c</sup> |
| HLA II eplet mismatch sum, n | 22 (8-28) | 23 (14-30) | 8 (7-25) | 0.269<sup>c</sup> |

HLA, human leukocyte antigen; IgA, immunoglobulin A; IQR, interquartile range; PRA, panel-reactive antibody.
<sup>a</sup>The significance of variation was calculated between rejection and non-rejection group of patients with rs7542235 GG-genotype.
<sup>b</sup>The medians of PRA values are calculated only from patients with PRA I >0 and PRA II >0.
<sup>c</sup>The Mann–Whitney U-test.
<sup>d</sup>The Pearson chi-square test.
<sup>e</sup>Fisher's exact test.
<sup>f</sup>The significance of variation was not analyzed due to the low frequency count.
<sup>g</sup> $n = 2$
<sup>h</sup> $n = 1$
<sup>f</sup>The Student's t-test.

DNA samples were extracted from whole blood at the time of histocompatibility testing for transplantation, and serum samples were collected for complement-dependent cytotoxic crossmatch to detect alloantibodies before and after transplantation at the Finnish Red Cross Blood Service, Helsinki, Finland. The clinical data were extracted from the Finnish Transplant Registry, which is a national follow-up registry obliged by law (**Table 1**).

### Genotyping and imputation

Genotyping and imputation procedures are explained in more detail in Markkinen et al. (6). Briefly, the genotyping was performed at the Finnish Institute of Molecular Medicine (FIMM), Helsinki, Finland using Illumina’s Infinium Global Screening array-24 v2.0 with multi-disease drop-in. The genotyped data were imputed with Finnish SISu v3 reference panel consisting of high-coverage WGS data from THL Biobank cohorts (N=1768) (Pärn et al. manuscript in preparation). The variant rs7542235 tagging for *CFHR*-deletion was imputed and had an estimated quality measurement value, INFO-score, of 0.98. The other variant investigated, rs6677604 tagging for *CFHR*-deletion was also imputed and had an INFO-score of 0.99.

### Multiplex ligation-dependent probe amplification

MLPA was performed for 15 homozygous rs7542235-G patients (eight with rejection, seven without rejection) to screen the copy number variations in *CFH/CFHR* genomic region. Additionally, we performed the analysis for control patients including eight individuals with heterozygous AG-genotype (four with rejection, four without rejection) and eight individuals with homozygous AA-genotype (four with rejection, four without rejection). The detailed description of MLPA procedure is provided in the **Supplementary data**.

### Whole genome sequencing

WGS was previously performed for three rs7542235 GG patients with rejection. The results and detailed information about the WGS for these samples are provided in the main text and supplementary data of our previous publication (6). Here, we sequenced seven more patients with GG-genotype (five with rejection, two without rejection). Sequencing was performed at FIMM, Helsinki, Finland, and the samples were sequenced with Illumina’s NovaSeq S4 NS4-300 run. The number of aligned bases for each sample was 20–50 Gb. The input reads for each sample were around 200–500 million, and the length of reads was 151 base pairs. The coverage depth for the sequenced samples was 7–17x. More detailed statistics for the WGS run are presented in **Supplementary Table S2**.

### Western blot for FH and FHR proteins

In total of 15 serum samples from nine rs7542235 GG patients (five with rejection, four without rejection) were available for western blot analysis of FH and FHR-1 expression. From the 15 serum samples, nine were collected before transplantation (pre-tx) and six were collected after transplantation (post-tx). The detailed methods for western blot are provided in the **Supplementary data**.

### Expression quantitative trait loci (eQTL) analyses

In the eQTL analyses, we used three databases to see the impact of Δ*CFHR3*–1 deletion tagging variant rs7542235 to gene expression: 1) The FIVEx browser (https://fivex.sph.umich.edu/) eQTL data from blood vessel and kidney samples from the EBI eQTL catalogue (4); 2) the eQTLGen Consortium database (https://www.eqtlgen.org/) including cis-eQTL, trans-eQTL and eQTS (associations between polygenic scores and gene expression levels) data from 37 datasets (3) from blood samples; and 3) the Human Kidney eQTL Atlas (https://susztaklab.com/Kidney_eQTL/index.php) including four studies with total of 686 microdissected human kidney tubule samples (5). Each of these three databases share data from the Genotype-Tissue Expression Consortium (GTEx). We used the same databases to also see the impact of an additional *ΔCFHR3–1* deletion tagging variant rs6677604 on gene expression. All the results are included regardless of P-value.

### Protein quantitative trait loci (pQTL), gene ontology and Reactome analyses

We used Olink (n = 1225) and SomaScan (n = 865) pQTL plasma proteomics data of healthy blood donors from FinnGen project (https://www.finngen.fi/en) to see whether the ΔCFHR311 deletion tagging variants rs7542235 and rs6677604 have effect on protein expression. Results with P-value of <3.49 x 10^-06^ were included. These genes were also included in the gene ontology (GO) enrichment analysis (https://geneontology.org/) for each three aspects of biological process, molecular function and cellular component, and Reactome pathway database (https://reactome.org/).

### Other statistical analyses

Characteristics of patients in **Table 1** and **Table 2** were described by medians and interquartile ranges (IQRs) or ranges, and frequencies and percentages. The comparison between the two groups of patients with rs7542235 GG-genotype and patients with AA/AG-genotype (**Table 1**), and GG-genotype with rejection and GG-genotype without rejection (**Table 2**) were analyzed using the nonparametric Mann-Whitney U-test for non-normally distributed data (recipient age, follow-up time, PRA I and II, HLA eplet mismatch, HLA I eplet mismatch and HLA II eplet mismatch), Pearson’s chi-square test for recipient sex, Fisher’s exact test for primary diagnosis, or Student’s t-test for normally distributed data (cold ischemia). The significance of variation of PRA-values in **Table 2** was not analyzed due to the low frequency count. P-values <0.05 were considered statistically significant.

## RESULTS

### Characteristics of the study population

We previously reported genetic association between reduced rejection-free survival and ΔCFHR311-deletion tagging variant rs7542235 genotype GG (6). 15 out of 1025 patients were homozygous for this deletion-tagging variant, 4% belonging to rejection-group and 1% to non-rejection group. The characteristics of the study groups are shown in **Table 1** and **Table 2**. In total of 53% of rs7542235 GG patients had rejection, whereas the percentage among AG/AA patients was only 19 (P-value <0.001). No significant difference in characteristics between those with rejection and without rejection among patients with rs7522235 GG genotype were found (**Table 2**). There was no enrichment of diagnoses known to be linked to *CFHR* locus.

### Multiplex ligation-dependent probe amplification

MLPA confirmed that all 15 rs7542235 GG patients had homozygous deletions of different sizes at the *CFH/CFHR* locus, three different types are depicted in the **Figure 1**. The shared feature of all samples was a homozygous deletion of *CFHR1* gene, indicating that variant rs7542235 tags for deletion of that gene. Patients with deletion type 1 were also homozygous for A allele of the other variant rs6677604, while patients with deletion type 2 and 3 were heterozygous AG for that variant indicating that it tags for a deletion of both *CFHR3* and *CFHR1*.

**Figure 1.**
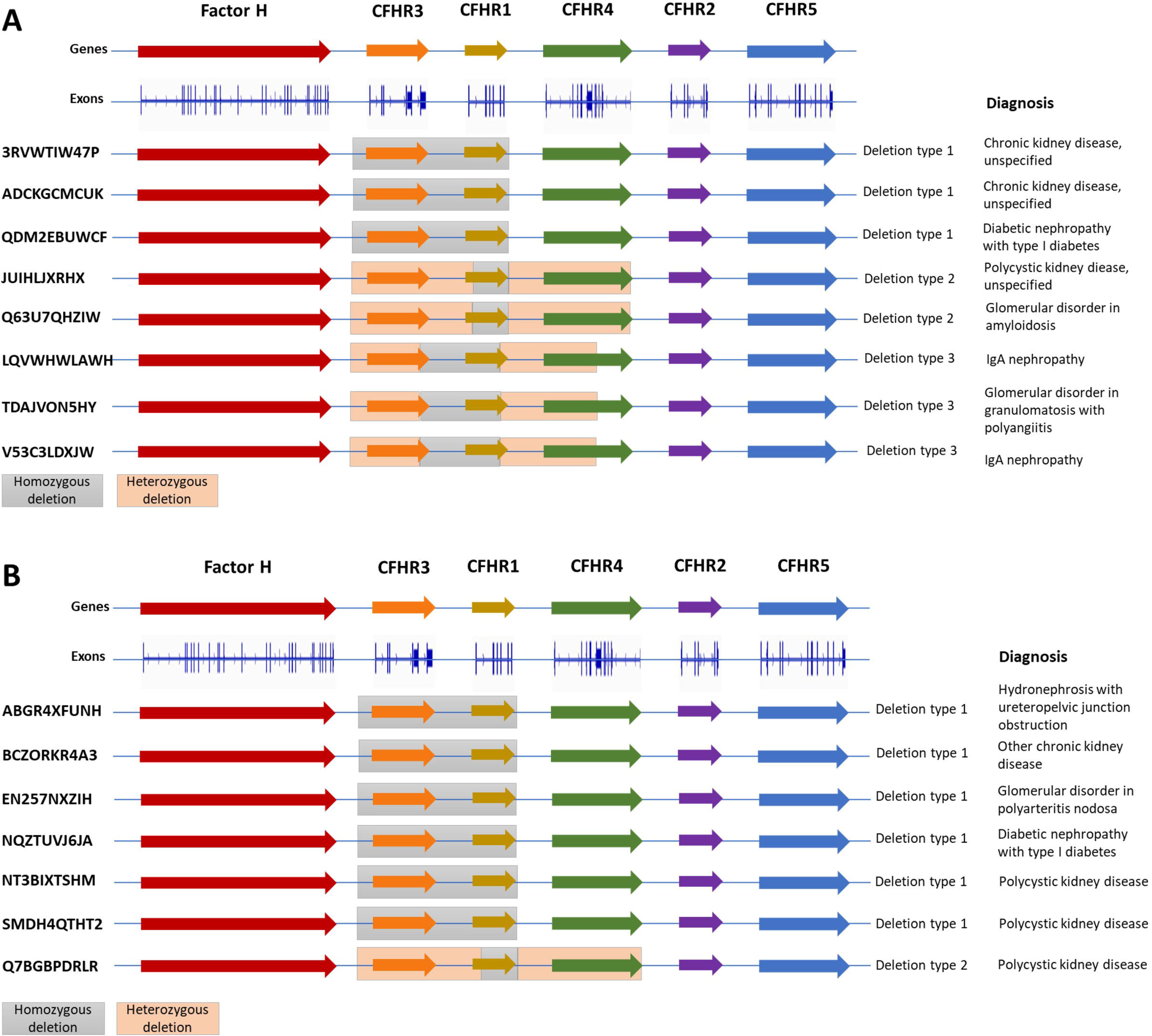
MLPA results for rs7542235 GG genotype patients with rejection (A) and without rejection (B). Deletion type 1 indicates homozygous deletion at *CFHR3* exons (ex) 1lll6 and *CFHR1* ex 1lll6. Deletion type 2 indicates homozygous deletion at *CFHR1* ex 2lll6 and heterozygous deletions at *CFHR3* ex 1lll6 and *CFHR4* ex 1lll10. Deletion type 3 indicates homozygous deletion at *CFHR3* ex 6 and *CFHR1* ex 1lll6 and heterozygous deletions at *CFHR3* ex 1lll4 and *CFHR4* ex 1lll6. CFHR, complement factor H related; MLPA, multiplex ligation-dependent probe amplification.

As assumed, eight patients with rs7542235 genotype AA had no deletions at the *CFHR* locus, and eight patients with heterozygous AG genotype carried a heterozygous deletion encompassing both the *CFHR1* and *CFHR3* genes (**Figure 2**). The patients with rs7542235 AA genotype were also homozygous GG for the variant rs6677604, while heterozygous rs7542235 AG patients were also heterozygous AG for variant rs6677604.

**Figure 2.**
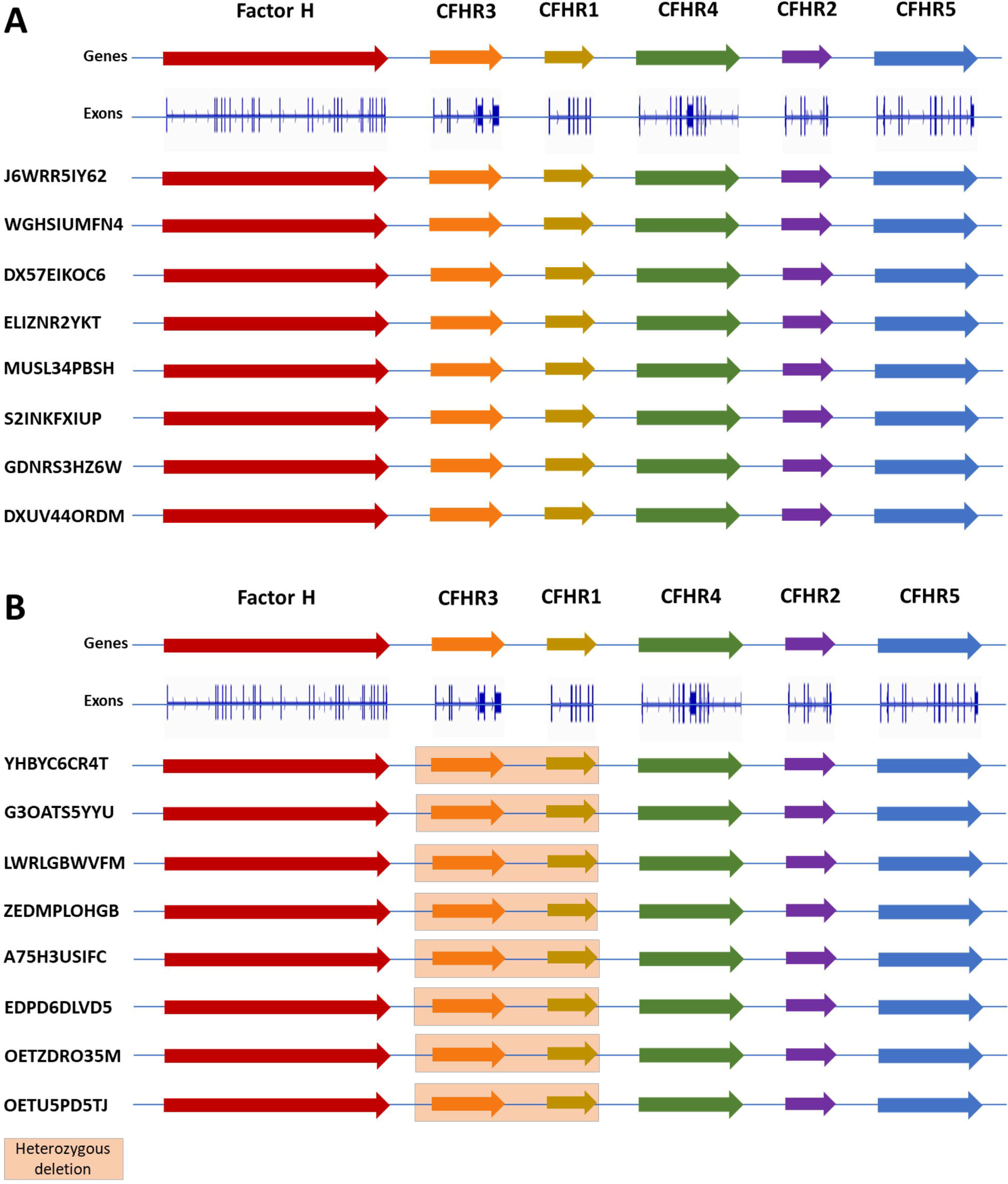
MLPA results for rs7542235 AA genotype (A) and AG genotype (B) patients. Patients with rs7542235 AA genotype had normal genotype without deletions. Patients with rs7542235 AG genotype carried a heterozygous deletion covering *CFHR3* ex 1lll6 and *CFHR1* ex 1lll6. CFHR, complement factor H related; MLPA, multiplex ligation-dependent probe amplification.

### Whole genome sequencing

WGS (**Figure 3**) confirmed the results of MLPA; all ten rs7542235 GG patients had homozygous deletions of different sizes but they shared the deletion of the *CFHR1* gene. In addition to homozygous deletions, WGS results confirmed the findings of MLPA results showing that 5 out of 10 patients carried heterozygous deletions. The **Figure 4** represents the deletion type 3 in more detail.

**Figure 3.**
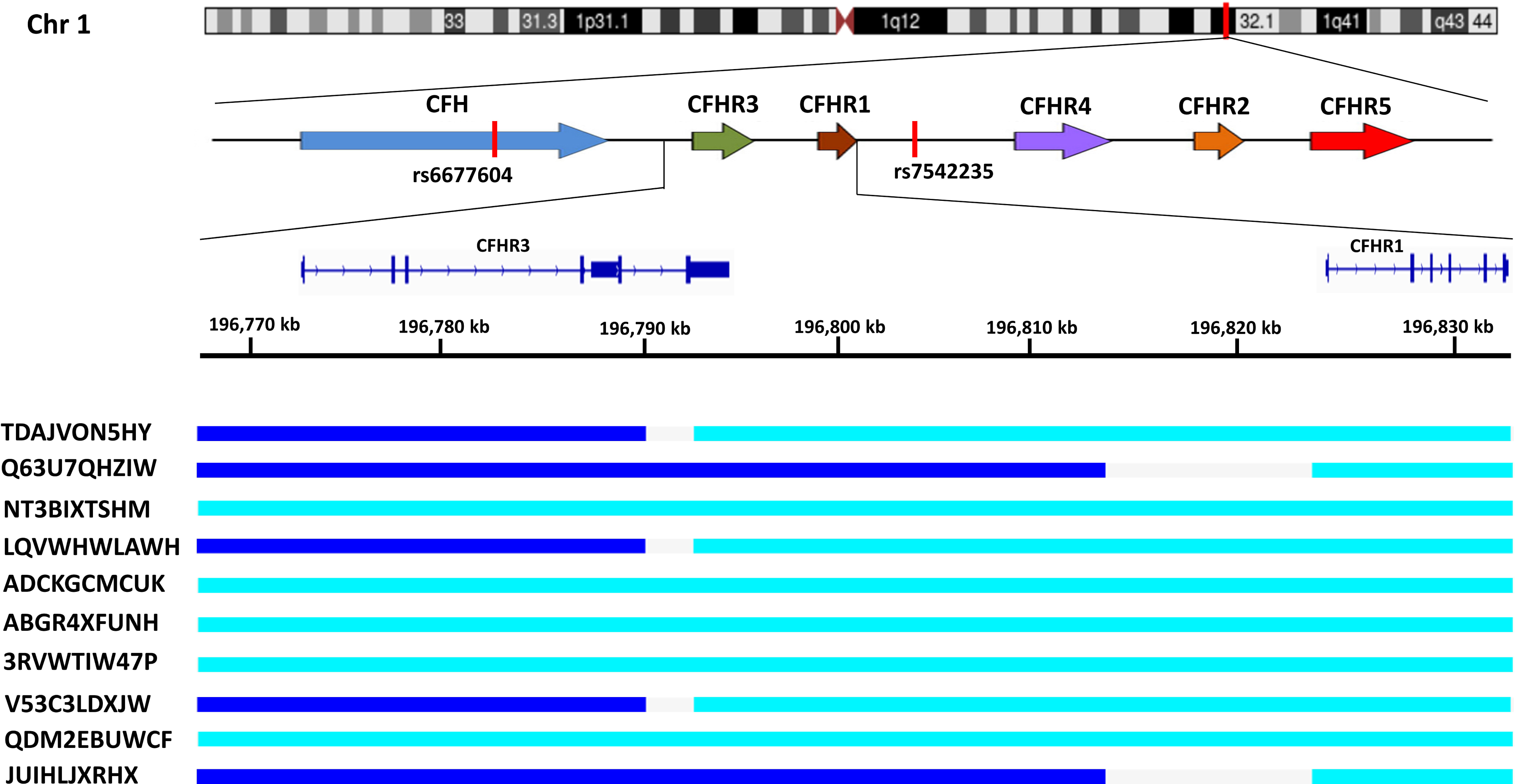
The results of whole genome sequencing of CFHR3fl1 loci on chromosome 1 for 10 rs7542235 GG genotype patients. The turquoise color represents a homozygous deletion, and the dark blue heterozygous deletion. The three lowest samples were sequenced at the time of Study I, other samples were sequenced at the time of Study II. The IDs on the left indicate the pseudonymes for each patient. The variant rs7542235 tagging for the deletion Δ*CFHR1* is located in the intergenic region between *CFHR1* and *CFHR4*, and the variant rs6677604 tagging for deletion ΔCFHR3fZ1 is located in the intron 11 of *CFH* gene (red bars). Each patient with homozygous deletion ΔCFHR3fZ1 were homozygous for rs6677604 AA genotype, while rest of the patients were heterozygous AG for that variant. CFH, complement factor H; CFHR, complement factor H related; ID, identity.

**Figure 4.**
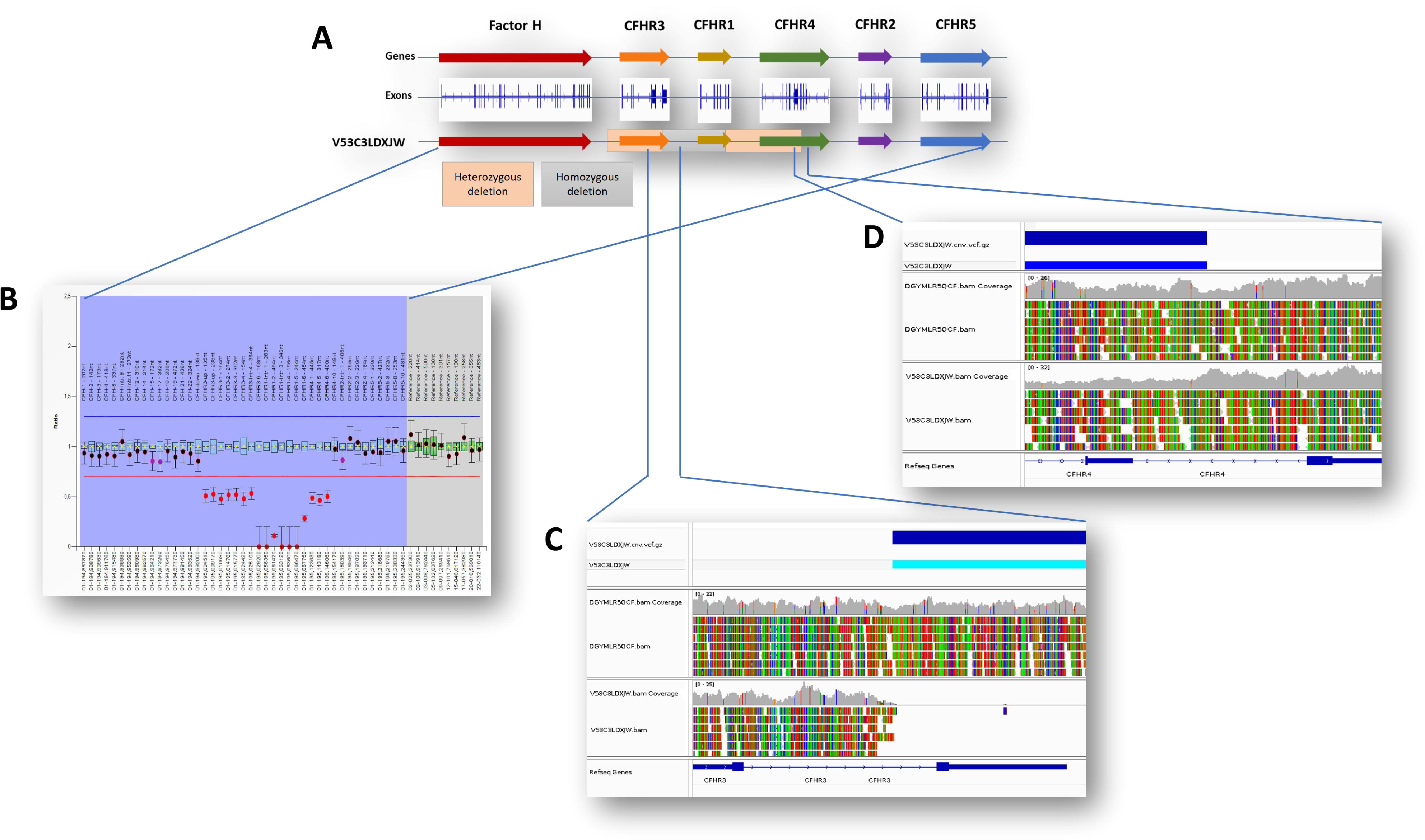
Detailed illustration of MLPA and WGS results for Deletion type 3. Patient with homozygous deletion covering *CFHR3* exon 6 and *CFHR1* exons 1lll6, and heterozygous deletions covering *CFHR3* exons 1lll4 and *CFHR4* exons 1lll4. A) *CFH/CFHR* gene locus, exons and deletions of patient V53C3LDXJW. B) MLPA result for the *CFH/CFHR* locus. The deletion is denoted by the red spots below the deletion cut-off line (red) in the ratio chart. The red line represents the 0.7 ratios, and the blue line represents the 1.3 ratios. Above the blue line are the genes covered by the SALSA MLPA probemix, P236 *CFH* Region B1. C) Visualization of homozygous deletion in WGS data at the beginning of *CFHR3* gene exon 6. Control sample without deletion above, the patient sample below. D) Visualization of heterozygous deletion in WGS data at the end of *CFHR4* gene. The coverage (gray) decreases half in size at the heterozygous deletion region. CFH, complement factor H; CFHR, complement factor H related; MLPA, multiplex ligation-dependent probe amplification; WGS, whole genome sequencing.

### Western blot of FH and FHR proteins

The western blot run showed that 9/9 of serum samples from individuals with rs7542235 GG genotype lacked the FHR-1 protein (**Figure 5**). The results are in line to those obtained from genetic MLPA and WGS analyses. The more detailed description and genotypes for each serum sample is shown in **Table 3**.

**Figure 5.**
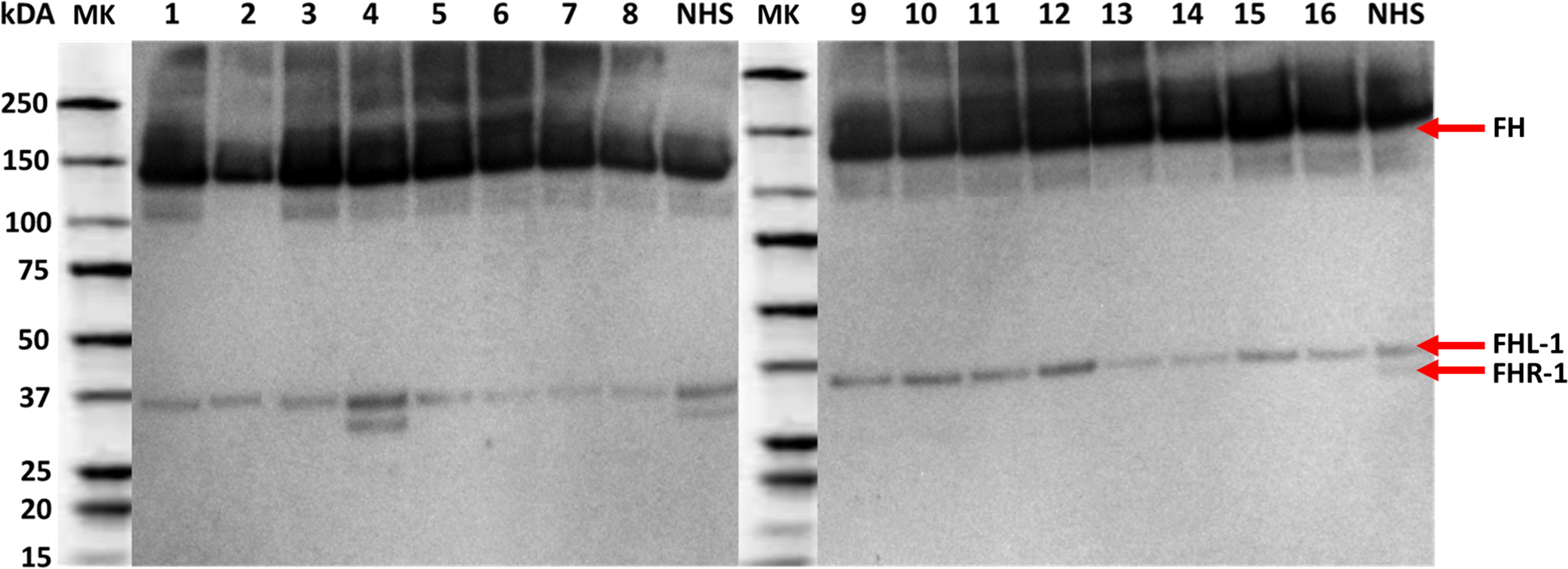
Western blot analyses of FH and FHR-1 protein expression. The detailed information for each serum sample is described in Table 3. The sample number 4 (control) and normal human serum (NHS, also control) have normal expression of FHR1. The other samples are individuals with homozygous deletion of *CFHR1* gene. FH, factor H protein; FHL-1, factor H like protein 1; FHR-1, factor H related protein 1; kDa, kilodalton; MK, marker; NHS, normal human serum

**Table 3.** Detailed description of serum samples.

| Sample ID<br>(pseudonymized) | Sample<br>number<br>in the<br>Figure 5 | Pre-tx<br>sample | Post-tx<br>sample | The region for<br>homozygous deletion<br>according to MLPA/WGS | The region for<br>heterozygous deletion<br>according to MLPA/WGS | Rejection* |
| --- | --- | --- | --- | --- | --- | --- |
| ABGR4XFUNH | 1 | x |  | CFHR3 (exons 1-6), CFHR1<br>(exons 1-6) |  | 0 |
| ABGR4XFUNH | 2 |  | x | CFHR3 (exons 1-6), CFHR1<br>(exons 1-6) |  | 0 |
| BCZORKR4A3 | 3 | x |  | CFHR3 (exons 1-6), CFHR1<br>(exons 1-6) |  | 0 |
| DGYMLR5QCF | 4 | x |  | no deletion control sample |  | 0 |
| EN257NXZIH | 5 | x |  | CFHR3 (exons 1-6), CFHR1<br>(exons 1-6) |  | 0 |
| NQZTUVJ6JA | 6 | x |  | CFHR3 (exons 1-6), CFHR1<br>(exons 1-6) |  | 0 |
| 3RVWTIW47P | 7 | x |  | CFHR3 (exons 1-6), CFHR1<br>(exons 1-6) |  | 1 |
| 3RVWTIW47P | 8 |  | x | CFHR3 (exons 1-6), CFHR1<br>(exons 1-6) |  | 1 |
| ADCKGCMCUK | 9 | x |  | CFHR3 (exons 1-6), CFHR1<br>(exons 1-6) |  | 1 |
| ADCKGCMCUK | 10 |  | x | CFHR3 (exons 1-6), CFHR1<br>(exons 1-6) |  | 1 |
| JUIHLJXRHX | 11 | x |  | CFHR1 (exons 1-6) | CFHR3 (exons 1-6), CFHR4<br>(exons 1-10) | 1 |
| JUIHLJXRHX | 12 |  | x | CFHR1 (exons 1-6) | CFHR3 (exons 1-6), CFHR4<br>(exons 1-10) | 1 |
| QDM2EBUWCF | 13 | x |  | CFHR3 (exons 1-6), CFHR1<br>(exons 1-6) |  | 1 |
| QDM2EBUWCF | 14 |  | x | CFHR3 (exons 1-6), CFHR1<br>(exons 1-6) |  | 1 |
| V53C3LDXJW | 15 | x |  | CFHR3 (exon 6), CFHR1<br>(exons 1-6) | CFHR3 (exons 1-4), CFHR4<br>(exons 1-6) | 1 |
| V53C3LDXJW | 16 |  | x | CFHR3 (exon 6), CFHR1<br>(exons 1-6) | CFHR3 (exons 1-4), CFHR4<br>(exons 1-6) | 1 |

CFHR, complement factor H related; MLPA, multiplex ligation-dependent probe amplification; WGS, whole genome sequencing; \*Acute rejection status 0 = no rejection, 1 = rejection

### Expression quantitative trait loci analyses

Both tagging variants are intronic, the rs7542235 residing in first intron of LOC100996886, a complement factor H-related protein 3-like pseudogene and rs6677604 residing 3’ downstream of exon 11 of *CFH*. To understand whether the ΔCFHR311 deletions or the intronic tagging variants rs7542235 and rs6677604 regulate the expression of other genes, we screened the eQTL databases for both variants. The results are presented in **Table 4**. According to FIVEx database, the allele G of rs7542235 and allele A of rs6677604 were both associated with the lower expression levels of *CFHR1, –3,* and *-4* genes. Additionally, the variant rs6677604 was also associated with increased expression level of *CFH* and lower expression of *KCNT2*. All the results from FIVEx were from GTEx study, including data from blood vessel and kidney samples. Based on the eQTLGen Consortium database consisting of only blood samples, both variants were associated merely with the higher expression level of *CFH*. When screening the Human Kidney eQTL Atlas having samples from kidney tubule, we found two statistically significant associations in *CFHR1* and *CFHR3* genes. The beta-values for these findings were negative indicating reduced expression.

**Table 4.**
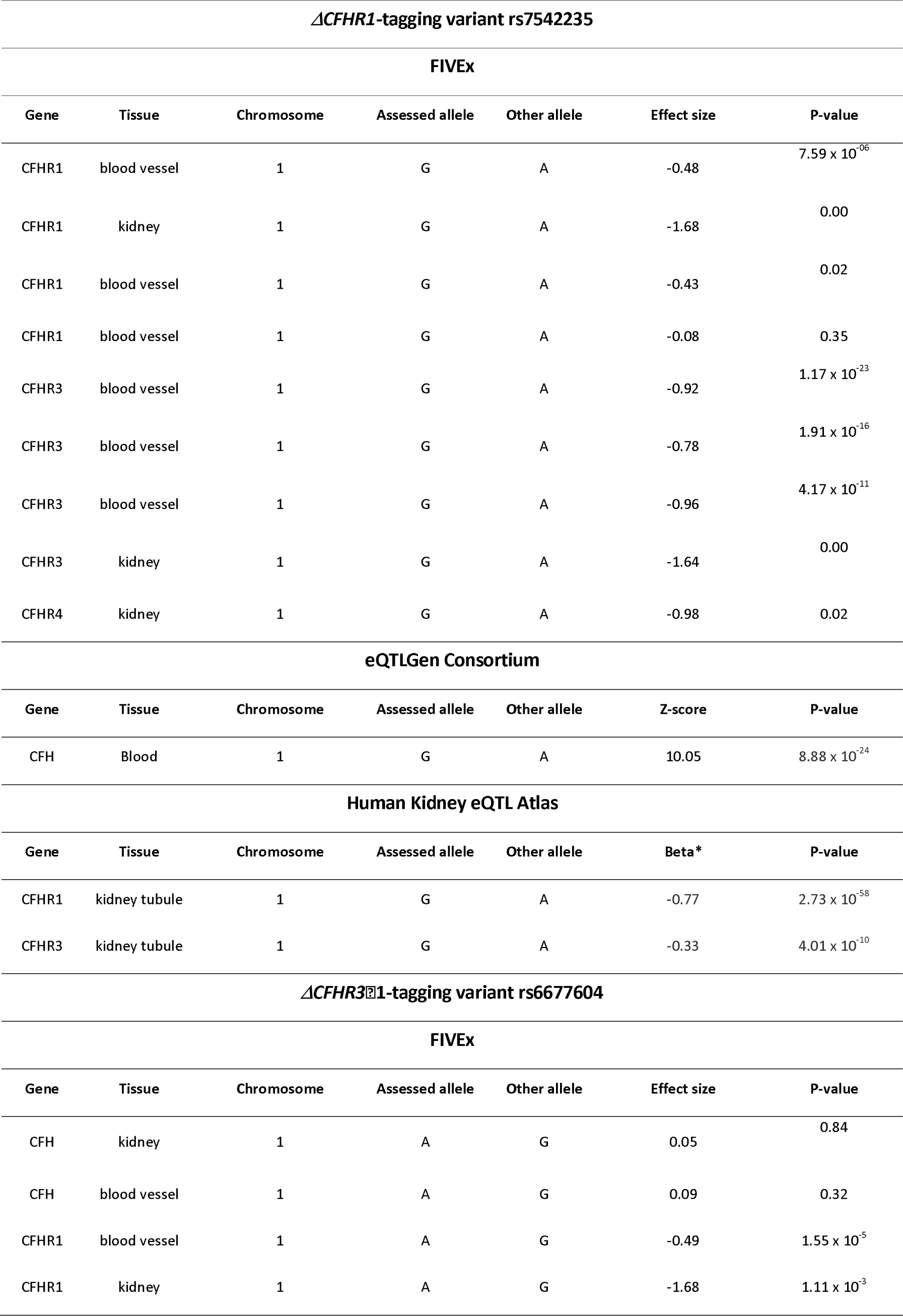
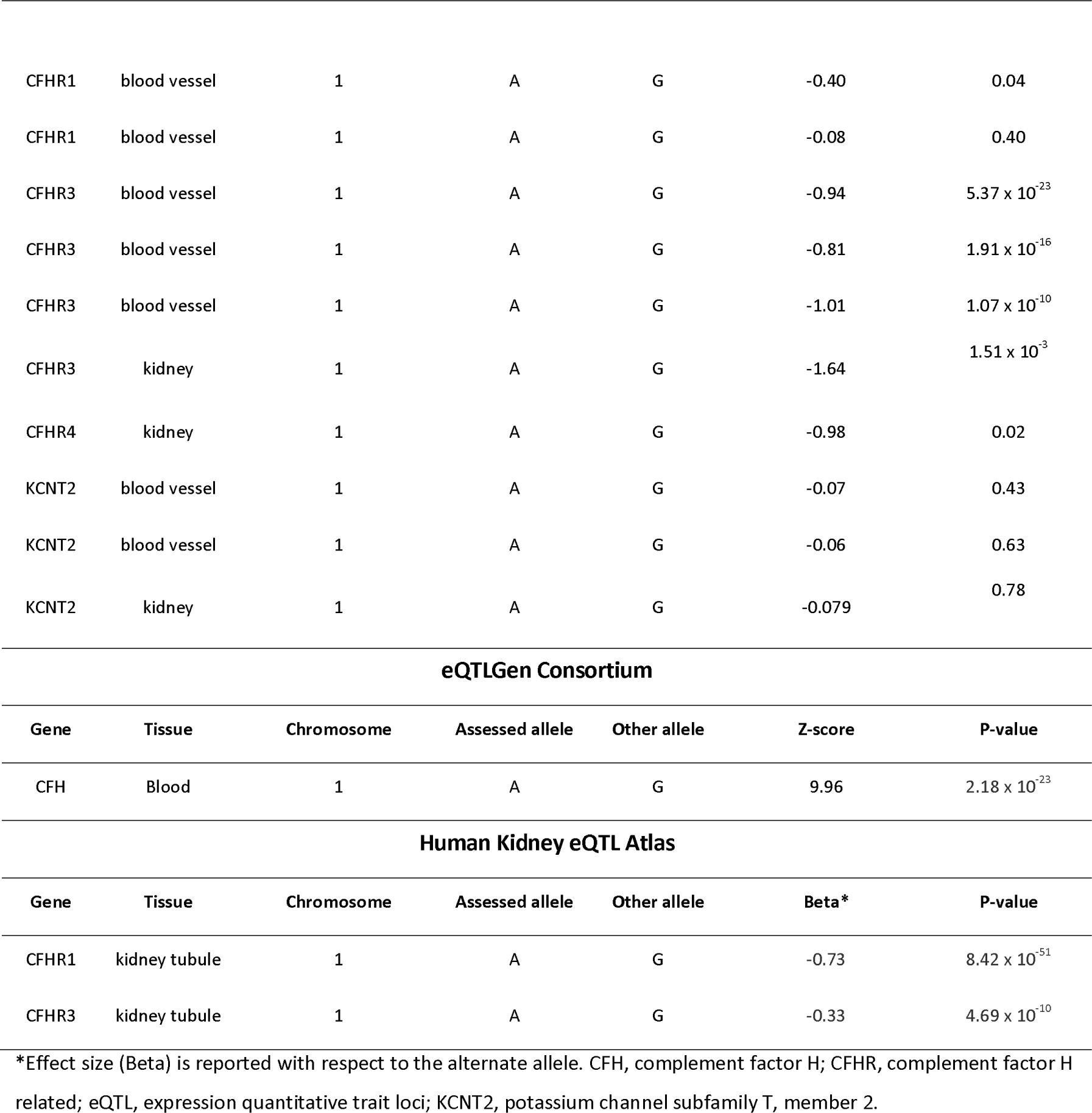
eQTL associations of CFHR3⍰1 deletion tagging variants rs7542235 and rs6677604.

### Protein quantitative trait loci, gene ontology and Reactome analyses

When analysing the effect of variant rs7542235 allele G on protein expression, total of 23 proteins were shown to be differentially expressed. The Olink results showed three genes to be associated with P-value <2.97 x 10^-8^; *CFH, CFHR2* and *CFHR5* (**Table 5**), each gene having a positive beta-value and thus meaning an increase in expression. SomaScan data resulted in total of 22 proteins to be differentially expressed with P-value <3.5 x 10^-6^. In concordance to Olink results, also SomaScan showed an increase in expression for *CFH* and *CFHR5*, and as was assumed, the Δ*CFHR1* deletion tagging variant resulted in decreased expression of *CFHR1* gene. Other differentially expressed genes for rs7542235 included, e.g., prolactin receptor (*PRLR*) with decreasing expression levels and Kirre Like Nephrin Family Adhesion Molecule 1 (*KIRREL1*) with increased expression. For variant rs6677604, the allele A was associated with two differentially expressed genes with P-value <2.87 x 10^-4^; CFH and Leukocyte Immunoglobulin Like Receptor A5 (LILRA5). For both genes, the beta-value was positive indicating increasing expression levels. Olink result for this variant resulted in five differentially expressed genes with P-value <1.78 x 10^-9^. Like variant rs7542235, also rs6677604 was associated with decreased level of *CFHR1*.

**Table 5.** pQTL associations of CFHR3⍰1 deletion tagging variants rs7542235 and rs6677604 in Olink and SomaScan data.

| *ΔCFHR1*-tagging variant rs7542235 |  |  |  |  |  |
| --- | --- | --- | --- | --- | --- |
| Olink (n = 1225) |  |  |  |  |  |
| Gene | Chromosome | Assessed allele | Other allele | Beta | P-value |
| CFH | 1 | G | A | 0.42 | $2.34 \times 10^{-12}$ |
| CFHR2 | 1 | G | A | 0.38 | $2.54 \times 10^{-10}$ |
| CFHR5 | 1 | G | A | 0.33 | $2.97 \times 10^{-08}$ |
| SomaScan (n = 865) |  |  |  |  |  |
| Gene | Chromosome | Assessed allele | Other allele | Beta | P-value |
| AKR1B10 | 7 | G | A | -0.87 | $6.01 \times 10^{-37}$ |
| CFH | 1 | G | A | 0.37 | $1.81 \times 10^{-07}$ |
| CFHR1 | 1 | G | A | -0.82 | $1.05 \times 10^{-33}$ |
| CFHR5 | 1 | G | A | 0.40 | $9.31 \times 10^{-09}$ |
| DYNLT3 | X | G | A | 0.37 | $1.95 \times 10^{-07}$ |
| F13B | 1 | G | A | 0.43 | $5.31 \times 10^{-10}$ |
| GLT8D1 | 3 | G | A | -0.41 | $5.62 \times 10^{-09}$ |
| KIRREL1 | 1 | G | A | 0.35 | $4.78 \times 10^{-07}$ |
| LRP1B | 2 | G | A | -0.34 | $9.86 \times 10^{-07}$ |
| MAN1B1 | 9 | G | A | 0.45 | $1.97 \times 10^{-10}$ |
| MYCT1 | 6 | G | A | -0.33 | $3.34 \times 10^{-06}$ |
| NAGLU | 17 | G | A | -0.63 | $1.11 \times 10^{-19}$ |
| NDUFS4 | 5 | G | A | 0.64 | $1.08 \times 10^{-20}$ |
| OTOR | 20 | G | A | -0.41 | $6.56 \times 10^{-09}$ |
| PRLR | 5 | G | A | -1.03 | $9.62 \times 10^{-55}$ |
| RIPPLY3 | 21 | G | A | 0.61 | $1.14 \times 10^{-18}$ |
| RPS6KA1 | 1 | G | A | -0.42 | $2.68 \times 10^{-09}$ |
| SCRN3 | 2 | G | A | 0.55 | $1.63 \times 10^{-15}$ |
| TST | 22 | G | A | 0.48 | $6.90 \times 10^{-12}$ |
| UGT2B15 | 4 | G | A | 0.56 | $1.25 \times 10^{-15}$ |
| ULBP2 | 6 | G | A | 0.44 | $3.29 \times 10^{-10}$ |
| ZDHHC4 | 7 | G | A | 0.33 | $3.50 \times 10^{-06}$ |
| *ΔCFHR3*1-tagging variant rs6677604 |  |  |  |  |  |
| Olink (n = 1225) |  |  |  |  |  |
| Gene | Chromosome | Assessed | Other allele | Beta | P-value |

| allele |  |  |  |  |  |
| --- | --- | --- | --- | --- | --- |
| CFH | 1 | G | A | 0.41 | $5.40 \times 10^{-17}$ |
| LILRA5 | 19 | G | A | 0.27 | $2.87 \times 10^{-8}$ |
| SomaScan (n = 865) |  |  |  |  |  |
| Gene | Chromosome | Assessed allele | Other allele | Beta | P-value |
| CFHR1 | 1 | G | A | -0.86 | $4.57 \times 10^{-36}$ |
| NAGLU | 17 | G | A | -0.64 | $5.89 \times 10^{-20}$ |
| OTOR | 20 | G | A | -0.43 | $1.78 \times 10^{-9}$ |
| PRLR | 5 | G | A | -1.03 | $3.04 \times 10^{-53}$ |
| RIPPLY3 | 21 | G | A | 0.63 | $3.48 \times 10^{-19}$ |

In the GO enrichment analysis of molecular function annotation, four out of 23 genes (*CFH*, *CFHR1*, *CFHR2* and *CFHR5*) for variant rs7542235 were associated with three functions of complement component C3b binding, opsonin binding and complement binding with false discovery rates (FDR) of 8.06 x 10^-6^, 3.71 x 10^-5^ and 5.34 x 10^-5^, respectively (**Supplementary table S4**). For the other variant rs6677604, GO enrichment analysis annotation showed an association of two out of seven genes (*CFH* and *CFHR1*) for complement component C3b binding pathway with FDR 2.76 x 10^-2^. The Reactome analysis showed that four out of 23 genes (*CFH, CFHR1, CFHR2* and *CFHR5*) were associated to two pathways; regulation of complement cascade and complement cascade with FDR 1.00 x 10^-3^ for both. For variant rs6677604, there were three associations; *CFH* and *CFHR1* were associated to regulation of complement cascade and complement cascade with FDR 9.04 x 10^-4^ for both, and *NAGLU* was associated to Sanfilippo syndrome with FDR 0.004. (**Supplementary table S4**).

## DISCUSSION

We reported earlier that homozygosity for variant rs7542235 allele G, tagging deletions at the *CFHR3–1* locus, predisposed patients to kidney allograft rejection. To understand better the mechanisms behind the association, we here fine-mapped the deletion boundaries and studied the effects of the gene deletion on protein levels. In addition to our previously reported variant, we also investigated another variant, rs6677604, known to tag for deletion at the *CFHR3–1* locus. Our results show that kidney transplantation patients with GG genotype of deletion-tagging variant rs7542235 have gene deletions of various sizes and types in the *CFHR* locus. The different deletion types, however, shared the complete deletion of the *CFHR1* gene pointing to its primary role. Also, we found that patients with homozygous ΔCFHR311 deletion are homozygous for rs6677604-A genotype. Patients with other types of deletion combinations (Deletion type 2 and 3, **Figure 1**) were heterozygous AG for rs6677604 indicating that allele A tags for a deletion of both *CFHR1* and *CFHR3*. With the support of our results and previous findings (20,21), it is plausible to assume that patients with deletion type 2 carry two heterozygous deletions of ΔCFHR311 and ΔCFHR114. Patients with deletion type 3, in addition to carrying a heterozygous deletion Δ*CFHR3-CFHR1*, consistently with previous studies, are likely to carry a heterozygous CFHR3::CFHR4 hybrid gene (22,23). The lack of expressed FHR-1 protein in homozygous rs7542235-G individuals was confirmed by Western blot analysis. We also showed by using both eQTL databases and proteomics studies that the rs7542235 allele G and rs6677604 allele A are associated with differences in plasma expression levels of various proteins, not merely with those encoded by genes at the *CFH/CFHR* locus but others as well.

The spectrum of kidney diseases in patients with homozygous *CFHR1* deletion was typical to adults receiving a kidney transplant in Finland. It did not involve complement-mediated diseases. Two patients carrying the deletion type 3 were diagnosed with IgA nephropathy and one with glomerular disorder in granulomatosis with polyangiitis. Previously, the CFHR3::CFHR4 hybrid gene has been observed in C3G (22) and atypical haemolytic uremic syndrome (23). This suggests that the polymorphism may in different patients result in varying manifestations of kidney disease. Similarly, other patients with the homozygous *CFHR1* deletion did not share any particular type of kidney disease.

Sequence and copy number variations in the *CFH a*nd *CFHR1–5* gene cluster have been linked to the kidney disorders such as C3 glomerulopathy (12), and IgA nephropathy (15). It has been shown that homozygous deletion Δ*CFHR3–1* and deficiency of FHR-1 and –3 have a protective effect in IgA nephropathy (24) and age-related macular degeneration (AMD) (25). A hybrid form of the *CFHR3–1* gene has been associated with an increased risk for C3 glomerulopathy (12). This finding indicates that an overactivity of FHRs is linked to complement-mediated diseases. The increased activity of FHRs is assumed to be due to a tendency to make dimers that more efficiently compete with FH. Only an autoimmune form of haemolytic uremic syndrome, with autoantibodies against FH, is linked to CFHR3–1 deficiency.

The capillaries of kidney glomerulus are continually perfused by large volumes of blood, so the FH and FHR complement proteins present in plasma are in direct contact with the membrane. Due to filtration of water into urine, the concentration of complement proteins increases as plasma flows through the kidney, thus exposing these glomerular cells and surfaces to higher concentrations of FH/FHR proteins than are found elsewhere in the body. Furthermore, the protection against spontaneous complement attack on the glomerulus relies exclusively on the effect of the soluble protector FH, whereas cellular surfaces of the body are further protected by surface bound complement regulators including membrane cofactor protein (MCP, CD46), decay accelerating factor (DAF, CD55) and protectin (CD59). These factors could explain why kidney is relatively sensitive to complement dysregulation (26).

A recent study by Sun *et al*. discovered that a mismatch in *LIMS1* was associated with death-censored graft loss and are expression quantitative trait loci in immune cells for *GCC2*, a gene encoding for a protein involved in mannose-6-phosphase receptor recycling and thus possibly triggering immune responses in the recipient (27). As shown by the eQTL and pQTL results, it is plausible that the homozygous deletion of *CFHR1* shared by all patients influences the expression levels of other *CFHR* genes and proteins, and thus could alter the complement activation. Among the FHR proteins, FHR-1 is most abundant in plasma with a typical molar ratio of the FHR-1 dimer to FH being approximately 0.3:1. The variant rs7542235 allele G is associated with an increased expression of FH and decreased expression of FHR-1. While FH inhibits complement activation and protects human endothelial cells, FHR-1 has an opposite effect. FH is a regulator of the C3 convertase and promotes degradation of C3b and opsonization of foreign pathogens with iC3b that results in phagocytosis. FHR-1 competes with FH for these activities, thereby increasing complement activation. It thus indirectly promotes C3/C5 convertase activity and complement membrane attack complex assembly. By inhibiting FH binding to C3b, FHR-1 promotes C5a formation and consequent inflammation. In the absence of FHR-1 and FHR-3, local FH binding and activity would be increased, resulting in suppression of complement activation, but, on the other hand, possibly also to an impaired ability to handle injured tissue components. Additionally, the disruption of the balance of FH/FHR protein concentrations and FHR-1 and –5 dimerization patterns due to genetic deletions and emergence of hybrid genes likely lowers the threshold of adverse immunological events in the presence of an additional triggering event such as organ transplant.

Besides *CFH/CFHRs,* differentially expressed genes based on proteomics data included *PRLR* and *KIRREL1* for variant rs7542235 and *PRLR* and *LILRA5* for rs6677604. The downregulated prolactin (PRL) is an endocrine hormone that has several physiological effects including immune system regulation and anti-inflammatory effects. It can trigger the production of proinflammatory cytokines, also having several anti-inflammatory effects that can reduce hyperinflammation (28). Prolactin exerts its effects through PRL receptors (PRLRs). The upregulated *KIRREL1,* also known as *NEPH1*, is a member of the NEPH protein family and is expressed in kidney podocytes, cells involved in ensuring size– and charge-selective ultrafiltration. Mutations in *KIRREL1* gene have been linked to hereditary nephrotic syndrome (29). Leukocyte immunoglobulin-like receptors (LILRs) are a family of inhibitory or stimulatory receptors expressed by immune cells, and several members of the LILR family recognize major histocompatibility complex class I (30). The function of the upregulated LILRA5 is currently unknown, but it has been shown to induce secretion of several proinflammatory cytokines, which suggests the roles of this protein in triggering innate immune response (31,32). The increased levels of LILRA5 have also been linked to antibody-mediated rejection after kidney transplantation (30) Overall, the eQTL analysis indicates a potentially enhanced inflammatory response in the presence of the tagging variants.

In conclusion, we found that rs7542235 allele G tags a deletion of the *CFHR1* gene. We also found that rs6677604 allele A tags for a deletion of the whole CFHR3fZ1 locus. The plasma proteomics studies also show that both variants are associated with an altered expression of FH/FHR proteins thus revealing a novel level of intricate regulation of complement system by the genetic polymorphisms of the *CFH* region.

## AUTHOR CONTRIBUTIONS STATEMENT

S. Markkinen, J. Partanen and K. Hyvärinen planned the study, interpreted the results and drafted the manuscript; S. Markkinen, I. Lokki and K. Hyvärinen carried out the genome data analyses; the MLPA and western blot analyses and interpretation of the results of these methods were carried out by I. Lokki, S. Markkinen and S. Meri; S. Markkinen executed *in silico* database searches; J. Ritari combined the pQTL summary statistics; I. Helanterä provided clinical data and expertise; all authors contributed to the final version of the manuscript.

## ACKNOWLEDGEMENT

We want to thank Maria Semenova, BSc, for help in collecting the serum samples. We also want to thank Lauri Snellman, biotechnology engineer, for help in laboratory with MLPA, and Marcel Messing, PhD, for help with the western blot method. The THL Biobank’s SISu v3 Imputation reference panel used for the research were obtained from THL Biobank (study number: BB2019_12). We thank all study participants for their generous participation in biobank research. We also want to thank the Sequencing Informatics Team, FIMM Human Genomics, University of Helsinki for the WGS analyses. We want to acknowledge the participants and investigators of FinnGen study. The FinnGen project is funded by two grants from Business Finland (HUS 4685/31/2016 and UH 4386/31/2016) and the following industry partners: AbbVie Inc., AstraZeneca UK Ltd, Biogen MA Inc., Bristol Myers Squibb (and Celgene Corporation & Celgene International II Sàrl), Genentech Inc., Merck Sharp & Dohme LCC, Pfizer Inc., GlaxoSmithKline Intellectual Property Development Ltd., Sanofi US Services Inc., Maze Therapeutics Inc., Janssen Biotech Inc, Novartis Pharma AG, and Boehringer Ingelheim International GmbH. Following biobanks are acknowledged for delivering biobank samples to FinnGen: Auria Biobank (www.auria.fi/biopankki), THL Biobank (www.thl.fi/biobank), Helsinki Biobank (www.helsinginbiopankki.fi), Biobank Borealis of Northern Finland (https://www.ppshp.fi/Tutkimus-ja-opetus/Biopankki/Pages/Biobank-Borealis-briefly-in-English.aspx), Finnish Clinical Biobank Tampere (www.tays.fi/en-US/Research_and_development/Finnish_Clinical_Biobank_Tampere), Biobank of Eastern Finland (www.ita-suomenbiopankki.fi/en), Central Finland Biobank (www.ksshp.fi/fi-FI/Potilaalle/Biopankki), Finnish Red Cross Blood Service Biobank (www.veripalvelu.fi/verenluovutus/biopankkitoiminta), Terveystalo Biobank (www.terveystalo.com/fi/Yritystietoa/Terveystalo-Biopankki/Biopankki/) and Arctic Biobank (https://www.oulu.fi/en/university/faculties-and-units/faculty-medicine/northern-finland-birth-cohorts-and-arctic-biobank). All Finnish Biobanks are members of BBMRI.fi infrastructure (www.bbmri.fi). Finnish Biobank Cooperative –FINBB (https://finbb.fi/) is the coordinator of BBMRI-ERIC operations in Finland. The Finnish biobank data can be accessed through the Fingenious^®^ services (https://site.fingenious.fi/en/) managed by FINBB.

## ETHICS STATEMENT

The study conforms to the principles of the Declaration of Helsinki and has been approved by the ethics committee of Helsinki University Hospital (HUS/1873/2018) and the Finnish National Supervisory Authority for Welfare and Health (V/9161/2019).

## REFERENCES

1. Steers NJ, Li Y, Drace Z, D’Addario JA, Fischman C, Liu L, et al. Genomic Mismatch at LIMS1 Locus and Kidney Allograft Rejection. New England Journal of Medicine. 2019;380(20):1918–28.

2. Markkinen S, Helanterä I, Lauronen J, Lempinen M, Partanen J, Hyvärinen K. Mismatches in gene deletions and kidney-related proteins as candidates for histocompatibility factors in kidney transplantation. Kidney Int Rep. 2022;7(11):2484–94.

3. Võsa U, Claringbould A, Westra HJ, Bonder MJ, Deelen P, Zeng B, et al. Large-scale cis– and trans-eQTL analyses identify thousands of genetic loci and polygenic scores that regulate blood gene expression. Nat Genet. 2021 Sep 1;53(9):1300–10.

4. Kwong A, Boughton AP, Wang M, Vandehaar P, Boehnke M, Abecasis G, et al. FIVEx: an interactive eQTL browser across public datasets. Bioinformatics. 2022 Jan 15;38(2):559–61.

5. Liu H, Doke T, Guo D, Sheng X, Ma Z, Park J, et al. Epigenomic and transcriptomic analyses define core cell types, genes and targetable mechanisms for kidney disease. Nat Genet. 2022 Jul 1;54(7):950–62.

6. Markkinen S, Helanterä I, Lauronen J, Lempinen M, Partanen J, Hyvärinen K. Mismatches in gene deletions and kidney-related proteins as candidates for histocompatibility factors in kidney transplantation. Kidney Int Rep. 2022;7(11):2484–94.

7. Lucientes-Continente L, Márquez-Tirado B, Goicoechea de Jorge E. The Factor H protein family: The switchers of the complement alternative pathway. Vol. 313, Immunological Reviews. Immunol Rev; 2023. p. 25–45.

8. Rodríguez De Córdoba S, Esparza-Gordillo J, Goicoechea De Jorge E, Lopez-Trascasa M, Sánchez-Corral P. The human complement factor H: Functional roles, genetic variations and disease associations. Mol Immunol. 2004 Jun;41(4):355–67.

9. Meri S, Pangburn MK. Discrimination between activators and nonactivators of the alternative pathway of complement: regulation via a sialic acid/polyanion binding site on factor H. Proc Natl Acad Sci U S A. 1990;87(10):3982–6.

10. Meri S. Self-nonself discrimination by the complement system. FEBS Lett. 2016 Aug 1;590(15):2418– 34.

11. Närkiö-Mäkelä M, Hellwage J, Tahkokallio O, Meri S. Complement-regulator factor H and related proteins in otitis media with effusion. Clin Immunol. 2001;100(1):118–26.

12. Malik TH, Lavin PJ, De Jorge EG, Vernon KA, Rose KL, Patel MP, et al. A hybrid CFHR3-1 gene causes familial C3 glomerulopathy. J Am Soc Nephrol. 2012 Jul;23(7):1155–60.

13. Józsi M, Licht C, Strobel S, Zipfel SLH, Richter H, Heinen S, et al. Factor H autoantibodies in atypical hemolytic uremic syndrome correlate with CFHR1/CFHR3 deficiency. Blood. 2008;111(3):1512–4.

14. Moore I, Strain L, Pappworth I, Kavanagh D, Barlow PN, Herbert AP, et al. Association of factor H autoantibodies with deletions of CFHR1, CFHR3, CFHR4, and with mutations in CFH, CFI, CD46, and C3 in patients with atypical hemolytic uremic syndrome. Blood. 2010;115(2):379–87.

15. Gharavi AG, Kiryluk K, Choi M, Li Y, Hou P, Xie J, et al. Genome-wide association study identifies susceptibility loci for IgA nephropathy. Nat Genet. 2011 Feb;43(4):321–9.

16. Yeo SC, Liu X, Liew A. Complement factor H gene polymorphism rs6677604 and the risk, severity and progression of IgA nephropathy: A systematic review and meta-analysis. Nephrology (Carlton). 2018 Dec 1;23(12):1096–106.

17. Xie J, Kiryluk K, Li Y, Mladkova N, Zhu L, Hou P, et al. Fine mapping implicates a deletion of CFHR1 and CFHR3 in protection from IgA nephropathy in Han Chinese. Journal of the American Society of Nephrology. 2016;27(10):3187–94.

18. Tao J, Tan M, Li LL, Chu H, Song D, Tan Y, et al. Genetic Variant CFH rs6677604 Might Play a Protective Role in lupus Nephritis. Am J Med Sci. 2021 Mar 1;361(3):336–43.

19. Roufosse C, Simmonds N, Clahsen-Van Groningen M, Haas M, Henriksen KJ, Horsfield C, et al. A 2018 Reference Guide to the Banff Classification of Renal Allograft Pathology. Transplantation. 2018;102(11):1795–814.

20. Abarrategui-Garrido C, Martínez-Barricarte R, López-Trascasa M, Rodríguez De Córdoba S, Sánchez-Corral P. Characterization of complement factor H-related (CFHR) proteins in plasma reveals novel genetic variations of CFHR1 associated with atypical hemolytic uremic syndrome. Blood [Internet]. 2009 Nov 5 [cited 2024 Jun 19];114(19):4261–71. Available from: https://pubmed.ncbi.nlm.nih.gov/19745068/

21. Moore I, Strain L, Pappworth I, Kavanagh D, Barlow PN, Herbert AP, et al. Association of factor H autoantibodies with deletions of CFHR1, CFHR3, CFHR4, and with mutations in CFH, CFI, CD46, and C3 in patients with atypical hemolytic uremic syndrome. Blood. 2010;115(2):379–87.

22. Piras R, Breno M, Valoti E, Alberti M, Iatropoulos P, Mele C, et al. CFH and CFHR Copy Number Variations in C3 Glomerulopathy and Immune Complex-Mediated Membranoproliferative Glomerulonephritis. Front Genet [Internet]. 2021 Jun 11 [cited 2024 Jun 19];12. Available from: https://pubmed.ncbi.nlm.nih.gov/34211499/

23. Piras R, Valoti E, Alberti M, Bresin E, Mele C, Breno M, et al. CFH and CFHR structural variants in atypical Hemolytic Uremic Syndrome: Prevalence, genomic characterization and impact on outcome. Front Immunol [Internet]. 2023 Jan 30 [cited 2024 Jun 19];13. Available from: https://pubmed.ncbi.nlm.nih.gov/36793547/

24. Kiryluk K, Novak J. The genetics and immunobiology of IgA nephropathy. J Clin Invest. 2014 Jun 2;124(6):2325–32.

25. Hughes AE, Orr N, Esfandiary H, Diaz-Torres M, Goodship T, Chakravarthy U. A common CFH haplotype, with deletion of CFHR1 and CFHR3, is associated with lower risk of age-related macular degeneration. Nat Genet. 2006 Oct;38(10):1173–7.

26. Thurman JM, Harrison RA. The susceptibility of the kidney to alternative pathway activation-A hypothesis. Immunol Rev. 2023 Jan 1;313(1):327–38.

27. Sun Z, Zhang Z, Banu K, Gibson IW, Colvin RB, Yi Z, et al. Multiscale genetic architecture of donor-recipient differences reveals intronic LIMS1 mismatches associated with kidney transplant survival. J Clin Invest. 2023 Sep 7;

28. Rasmi Y, Jalali L, Khalid S, Shokati A, Tyagi P, Ozturk A, et al. The effects of prolactin on the immune system, its relationship with the severity of COVID-19, and its potential immunomodulatory therapeutic effect. Cytokine. 2023 Sep 1;169:156253.

29. Sellin L, Huber TB, Gerke P, Quack I, Pavenstädt H, Walz G. NEPH1 defines a novel family of podocin interacting proteins. The FASEB journal[]: official publication of the Federation of American Societies for Experimental Biology. 2003;17(1):115–7.

30. Lamarthée B, Genet C, Cattin F, Danger R, Giral M, Brouard S, et al. Single-cell mapping of leukocyte immunoglobulin-like receptors in kidney transplant rejection. Frontiers in Transplantation. 2022 Aug 11;1:952785.

31. Mitchell A, Rentero C, Endoh Y, Hsu K, Gaus K, Geczy C, et al. LILRA5 is expressed by synovial tissue macrophages in rheumatoid arthritis, selectively induces pro-inflammatory cytokines and IL-10 and is regulated by TNF-alpha, IL-10 and IFN-gamma. Eur J Immunol [Internet]. 2008 [cited 2024 Jun 19];38(12):3459–73. Available from: https://pubmed.ncbi.nlm.nih.gov/19009525/

32. Truong AD, Hong Y, Nguyen HT, Nguyen CT, Chu NT, Tran HTT, et al. Molecular identification and characterisation of a novel chicken leukocyte immunoglobulin-like receptor A5. Br Poult Sci [Internet]. 2021 [cited 2024 Jun 19];62(1):68–80. Available from: https://pubmed.ncbi.nlm.nih.gov/32812773/

