## Supplementary for "The impact of complement factor H-related protein gene deletions on kidney transplantation"

Running title: CFHR deletions in kidney transplantation

Markkinen Salla^1†^, Lokki A. Inkeri^2,3†^, Helanterä Ilkka^4^, Ritari Jarmo^1^, Partanen Jukka^1^, Meri Seppo^2,5^, Hyvärinen Kati^1*^

^1^Research and Development, Finnish Red Cross Blood Service, Helsinki Finland

^2^Translational Immunology Research Program and Department of Bacteriology and Immunology, Faculty of Medicine, University of Helsinki, Helsinki, Finland

^3^Heart and Lung Center, Helsinki University Hospital and University of Helsinki, Helsinki, Finland

^4^Transplantation and Liver Surgery, Helsinki University Hospital and University of Helsinki, Helsinki, Finland

^5^Diagnostic Center, Helsinki University Hospital, Helsinki, Finland

† These authors contributed equally to this work

Keywords: Complement factor H-related, CFHR, kidney transplantation, gene deletion, allograft rejection

Number of words: 3884

^*^Corresponding author

Kati Hyvärinen, PhD, Associate professor

Finnish Red Cross, Blood Service

Research and Development

Biomedicum Helsinki 1, Haartmaninkatu 8

00290 Helsinki, FINLAND

**Table of contents**

Detailed methods	3

Multiplex ligation-dependent probe amplification (MLPA)	3

Western blot for CFH, CFHR1 and CFHR2 proteins	3

Supplementary tables	5

Supplementary table S1. P236-B1 MLPA probes arranged according to chromosomal location	5

Supplementary table S2. General statistics of the whole genome sequencing with Illumina’s NovaSeq S4 NS4-300 run	9

Supplementary table S3. WGS results. The exact positions of homozygous/heterozygous deletions	10

Supplementary table S4. Gene Ontology molecular functions and Reactome pathways associated with the genes from pQTL data	11

DETAILED METHODS

*Multiplex ligation-dependent probe amplification (MLPA)*

SALSA MLPA kit P236-A3 and SALSA MLPA Probemix P236-B1 (MRC Holland) containing 53 probes for CFH/CFHR genes (including 16 probes for *CFH*, 8 probes for *CFHR3*, 6 probes for *CFHR1*, 4 probes for *CFHR4*, 4 probes for *CFHR2*, and 5 probes for *CFHR5* (Supplementary table 1) were used. In the MLPA reaction, 5 μl of the DNA (15 ng/ul) per each sample were used and three normal control samples were included. The samples were diluted in low EDTA TE-buffer (10 mM Tris, pH 8.0, and 0.1 mM EDTA). First, the DNA was denatured 5 minutes at 98 °C with pause at 25 °C. After denaturation, the single-stranded DNAs were incubated with the probes in the hybridization reaction for 1 minute at 95 °C and 16-20 hours at 60 °C. After hybridization, the probes were ligated with ligation reaction; pause at 54 °C, 15 minutes at 54 °C, 5 minutes at 98 °C and finally pause at 20 °C. The ligated probes were then amplified by polymerase chain reaction (PCR) for 35 cycles (30 seconds at 95 °C, 30 seconds at 60 °C and 60 seconds at 72 °C) and finally incubated 20 minutes at 72 °C and paused at 15 °C. MiniAmp thermal cycler (Applied biosystems MiniAmp Plus) was used for the reaction.

After PCR reaction, 2 ul of the FAM-MLPA PCR product and 10 ul of HiDi Formamide (Applied Biosystems) were transferred to FIMM for separation by capillary electrophoresis (ABI3730xl DNA Analyzer). The sizes of the amplified fragments were identified according to their migration relative to the GeneScan™ 500 LIZ™ Dye Size Standard (Applied Biosystems). The amount of PCR products were determined with Coffalyser provided online by the manufacturer ([www.mlpa.com](http://www.mlpa.com)).

*Western blot for CFH and CFHR1 proteins*

Sera were diluted 1/100 in PBS and mixed with 4x Bolt LDS Sample Buffer (Invitrogen #B0007) to a concentration of 1x LDS. 20 µl of each sample were loaded on a Bolt 4-12% Bis-Tris Plus gel (Invitrogen #NW0412BOX) in Bolt MES SDS Running Buffer (Invitrogen #B0002) and ran for 45 minutes at 165 V. The proteins were transferred to nitrocellulose using iBlot 2NC Regular Stacks (Invitrogen #IB23001) in the iBlot 2 Gel Transfer Device (Invitrogen #IB21001) at 20 V for 1 min, then 23 V for 4 min, then 25 V for 2 min. The blots were blocked in glass bowls with 10 ml PBS / Tween‑20 0.05% / milk 5% for 1 h at RT on a rotating table. Goat-anti-human factor H (Calbiochem, Temecula, CA, USA) was added to a dilution of 1/10,000 in PBS / Tween‑20 0.05% / milk 5% and the blots were incubated o/n at 4 ^o^C on a rotating table. Then the blots were washed for 1 h at RT on a rotating table with several changes of PBS / Tween‑20 0.05%. Subsequently, the blots were incubated with polyclonal rabbit-anti-goat-HRP (Dako, Glostrup, Denmark) at 1/10,000 in PBS / Tween‑20 0.05% / milk 5% for 1 h at RT on a rotating table. After washing the blots as before, bands were made visible by an in-house method of enhanced chemiluminescence using Luminol and coumaric acid.

SUPPLEMENTARY TABLES

**Supplementary table S1. P236-B1 MLPA probes arranged according to chromosomal location.**

| **Length (nt)** | | **SALSA MLPA probe** | **Exon** | **Ligation site** | **Partial sequence (24 nt adjacent to ligation site)** | **Distance to next probe** |
| --- | --- | --- | --- | --- | --- | --- |
| **CFH gene (NM_000186.4)** | | | | | | |
|  | |  | start codon | 76-78 (exon 1) |  |  |
| 202 | | 07820-L07574 | exon 1 | 120-119 reverse | TCTGCTACACAA-ATAGCCCATAAC | 20.9 kb |
| 142 | | 07821-L07575 | exon 2 | 194-195 | CTGGTCTGACCA-AACATATCCAGA | 0.9 kb |
| 179 | | 07822-L07576 | exon 3 | 357-358 | ACTCCTTTTGGT-ACTTTTACCCTT | 2.1 kb |
| 419 | | 07823-L16758 | exon 4 | 458-459 | TAATTACCGTGA-ATGTGACACAGA | 3.7 kb |
| 337 | | 07824-L07578 | exon 6 | 818-819 | TGAAAGAGGAGA-TGCTGTATGCAC | 23.4 kb |
| 292 | | 22079-L31050 | intron 9 | 10.6 kb before exon 10 reverse | CAGAGCCACAGA-TAACAAGTCAGC | 13.7 kb |
| 373 | | 07827-L07582 | intron 11 | 1.1 kb after exon 11 | CTTGGACACATT-ATGATTGAGTCG | 8.4 kb |
| 310 | | 07828-L07583 | exon 12 | 1893-1894 | ATAGTTGGACCT-AATTCCGTTCAG | 1.6 kb |
| 214 | | 22074-L31045 | exon 14 | 2203-2202 reverse | ATCTCCATAGTA-ATAAGGAGGGGA | 1.6 kb |
| 172 | | 22071-L31042 | exon 15 | 2440-2441 | AAGGATGGATAC-ACACAGTCTGCA | 9.1 kb |
| 382 | | 07830-L07586 | exon 17 | 2719-2720 | ACGGAACCATTA-ATTCATCCAGGT | 3.2 kb |
| 208 | | 22073-L31044 | exon 18 | 2965-2966 | TTGAAGGTTTTG-GAATTGATGGGC | 1.3 kb |
| 472 | | 22559-L31056 | exon 19 | 3156-3157 | ATGGATGGAGCC-AGTAATGTAACA | 3.7 kb |
| 436 | | 22082-L31053 | exon 21 | 86 nt before exon 21 reverse | AACACAGCACTG-TATATAATATCA | 2.1 kb |
| 324 | | 22044-L31698 | exon 22 | 283 nt after exon 22 | TATCAATACATA-AATGCACCAAAA | 8.5 kb |
|  | |  | stop codon | 3769-3771 (Exon 22) |  |  |
| 139 | | 22043-L08618 | downstream | 8.8 kb after exon 22 | TGCACTTATACA-TGCAATCCGTTG | 12.5 kb |
| **CFHR3 gene (NM_021023.6)** | | | | | | |
| 135 | | 22996-L32432 | upstream | 6.1 kb before exon 1 | TTAGTCCGAGGT-AGAAAGGGACAT | 4.7 kb |
| 238 | | 22997-L32433 | upstream | 1.4 kb before exon 1 | GGGTGGTAATCT-TGGCTCTCAGTG | 1.5 kb |
|  | |  | start codon | 48-50 (exon 1) |  |  |
| 164 | | 07832-L07588 | exon 1 | 8 nt after exon 1 | CAAGGTAAGTTA-AAAGAGATCTAA | 4.1 kb |
| 274 | | 07833-L07589 | exon 2 | 100 nt before exon 2 | AACATTTTCTTG-TGGAATTACAGC | 1.0 kb |
| 392 | | 07834-L07590 | exon 3 | 61 bt after exon 3 | CACGGACGACAG-TCTCAGACTTGT | 8.6 kb |
| 154 | | 22069-L31040 | exon 4 | 287 nt after exon 4 | ATCAGCAAAATA-TGTTAGTTGCCA | 0.7 kb |
| 364 | | 07835-L07591 | intron 4 | 680 nt before exon 5 | GGGGGTTATATG-AATTCCTACATT | 4.1 kb |
| 168 | | 08218-L09921 | exon 6 | 1005-1004 reverse | TATCCCTTCCCG-ACACACTGCTTG | 27.2 kb |
|  | |  | stop codon | 1038-1040 (exon 6) |  |  |
| **CFHR1 gene (NM_002113.3)** | | | | | | |
|  | |  | start codon | 115-117 (exon 1) |  |  |
| 283 | | 22112-L31100 | intron 1 | 730 nt after exon 1 | TATGTCTGTACA-TGGAGTTTCGAT | 5.1 kb |
| 494 | | 22087-L31058 | exon 2 | 12 nt after exon 2 reverse | TTAATGAACAGA-GCATTTACTCAC | 1.7 kb |
| 346 | | 07839-L07595 | intron 3 | 396 nt after exon 3 reverse | AGAGAGTTTCAG-GTCCATGTGTAG | 0.5 kb |
| 196 | | 22072-L31043 | exon 4 | 194 nt before exon 4 reverse | CAGGTACAAGCT-TTGATGTTTTAA | 2.9 kb |
| 244 | | 22076-L31047 | exon 5 | 64 nt after exon 5 | ATTTTGCTGTTG-GTAACAAAATAA | 1.3 kb |
| 454 | | 22995-L32431 | exon 6 | 1120-1121 | ATCAATCATAAA-ATGCACACCTTT | 55.9 kb |
|  | |  | stop codon | 1834-1836 (exon 10) |  |  |
| **CFHR4 gene (NM_001201550.3)** | | | | | | |
|  | |  | start codon | 100-102 (exon 1) |  |  |
| 445 | | 22084-L31055 | exon 1 | 135 nt before exon 1 reverse | CTGGTATGTACA-TGTACAGCTTTA | 19.6 kb |
| 317 | | 2294-L32539 | exon 5 | 849-850 | CAGGGTTCTAAA-TATGTAACATGT | 2.9 kb |
| 400 | | 22558-L31052 | exon 6 | 938-939 | TCAACATGGACA-TCTATATTATGA | 8.1 kb |
| 148 | | 22111-L31098 | exon 10 | 1872-1873 | TGTCCAACTTCC-ACTTCTCACTCT | 55.9 kb |
|  |  |  | stop codon | 1834-1836 (exon 10) |  |  |
| **CFHR2 gene (NM_005666.4)** | | | | | | |
|  | |  | start codon | 144-146 (exon 1) |  |  |
| 406 | | 22113-L31101 | intron 1 | 729 nt after exon 1 reverse | TCGAAACTCCAA-GTACAGACATAG | 5.1 kb |
| 265 | | 07842-L07598 | exon 2 | 74 nt after exon 2 | CAAGATCATAAA-CACTTGATAATC | 1.6 kb |
| 226 | | 21368-L31327 | exon 3 | 268 nt after exon 3 | GTAATACCTGTG-TGTGGTTTATAG | 6.7 kb |
| 184 | | 07844-L07600 | exon 4 | 655-656 | ATATGCTCCAGG-TTCATCAGTTGA | 19.7 kb |
|  | |  | stop codon | 954-956 (exon 5) |  |  |
| **CFHR5 gene (NM_030787.4)** | | | | | | |
|  | |  | start codon | 110-112 (exon 1) |  |  |
| 330 | | 07845-L30998 | exon 1 | 152-153 | CATGGGTATCCA-CTGTTGGGGGAG | 5.2 kb |
| 427 | | 07846-L16757 | exon 2 | 223-224 | GATGAAGAAGAT-TATAACCCTTTT | 1.1 kb |
| 232 | | 07847-L07603 | exon 3 | 414-415 | ATCTTCAGGACT-AATACATCTGGA | 18.6 kb |
| 253 | | 22077-L31048 | exon 8 | 1380-1379 reverse | CTTTTGCTTCTG-GAAGTAGATAGT | 6.0 kb |
| 481 | | 22086-L31057 | exon 10 | 1762-1761 reverse | CGAAATGGTGGT-GATGATATCATC |  |
|  | |  | stop codon | 1817-1819 (exon 10) |  |  |

MLPA, multiplex ligation-dependent probe amplification; CFH, complement factor H; CFHR, complement factor H related.

**Supplementary table S2. General statistics of the whole genome sequencing with Illumina’s NovaSeq S4 NS4-300 run.**

| **Sample Name** | **Total number of variants** | **Sex** | **Total number of aligned reads** | **Total number of aligned bases** | **Coverage depth** | **% of sites in region with at least 20x coverage** | **Total number of input reads, millions** | **% of unmapped reads** | **% of properly paired reads** | **Median insert size** |
| --- | --- | --- | --- | --- | --- | --- | --- | --- | --- | --- |
| 3RVWTIW47P | 5161431 | XY | 262.8 | 38863.2 | 12.9 x | 94.4 | 407.8 | 0.5 | 97.7 | 453 |
| ABGR4XFUNH | 4893599 | XY | 352.1 | 52388.4 | 17.4 x | 94.5 | 423.9 | 0.6 | 97.5 | 439 |
| ADCKGCMCUK | 4711490 | XY | 271.2 | 40094.9 | 13.3 x | 94.4 | 494.3 | 0.6 | 97.4 | 460 |
| DGYMLR5QCF | 5924760 | XX | 273.8 | 40715.2 | 13.5 x | 94.1 | 236.6 | 0.5 | 97.7 | 424 |
| LQVWHWLAWH | 4869748 | XY | 320.4 | 47460.0 | 15.7 x | 94.3 | 389.6 | 0.6 | 97.5 | 393 |
| NT3BIXTSHM | 4389518 | XY | 150.0 | 22318.8 | 7.4 x | 93.0 | 561.5 | 0.5 | 97.8 | 467 |
| Q63U7QHZIW | 4431168 | XY | 155.0 | 22977.6 | 7.6 x | 93.3 | 415.7 | 0.6 | 97.4 | 489 |
| TDAJVON5HY | 4772389 | XY | 259.5 | 38188.8 | 12.7 x | 94.3 | 232.7 | 0.4 | 97.7 | 486 |

**Supplementary table S3. WGS results. The exact positions of homozygous/heterozygous deletions.**

| **Sample Name** | **Homozygous deletion, genes (exons)** | **Homozygous deletion, position** | **Heterozygous deletions, genes (exons)** | **Heterozygous deletions, positions** |
| --- | --- | --- | --- | --- |
| TDAJVON5HY | CFHR3 (exon 6), CFHR1 (exons 1─6) | chr1:196791783-196832580 | CFHR3 (exons 1─4), CFHR4 (exons 1─9) | chr1:196766991-196789398, chr1:196851189-196917464 |
| Q63U7QHZIW | CFHR1 (exons 1─6) | chr1:196822571-196832580 | CFHR3 (exons 1─6), CFHR4 (exons 1─10) | chr1:196766991-196812282, chr1:196851189-196933978 |
| NT3BIXTSHM | CFHR3 (exons 1─6), CFHR1 (exons 1─6) | chr1:196766991-196832580 | - | - |
| LQVWHWLAWH | CFHR3 (exon 6), CFHR1 (exons 1─6) | chr1:196791782-196832580 | CFHR3 (exons 1─4), CFHR4 (exons 1─9) | chr1:196766991-196789398, chr1:196851189-196917464 |
| DGYMLR5QCF | no deletion | no deletion | no deletion | no deletion |
| ADCKGCMCUK | CFHR3 (exons 1─6), CFHR1 (exons 1─6) | chr1:196766990-196832580 | - | - |
| ABGR4XFUNH | CFHR3 (exons 1─6), CFHR1 (exons 1─6) | chr1:196766991-196832580 | - | - |
| 3RVWTIW47P | CFHR3 (exons 1─6), CFHR1 (exons 1─6) | chr1:196766991-196832580 | - | - |
| V53C3LDXJW | CFHR3 (exon 6), CFHR1 (exons 1─6) | chr1:196792618-196833414 | CFHR3 (exons 1─4), CFHR4 (exons 1─9) | chr1:196767834-196788257, chr1:196851766-196913910 |
| QDM2EBUWCF | CFHR3 (exons 1─6), CFHR1 (exons 1─6) | chr1:196767834-196833414 | - | - |
| JUIHLJXRHX | CFHR1 (exons 1─6) | chr1:196817315-196833414 | CFHR3 (exons 1─6), CFHR4 (exons 1─10) | chr1:196767833-196812817, chr1:196851766-196934405 |

CFHR, complement factor H related; WGS, whole genome sequencing.

**Supplementary table S4. Gene Ontology molecular functions and Reactome pathways associated with the genes from pQTL data for variants rs7542235 and rs6677604.**

| **Variant rs7542235** | | | |
| --- | --- | --- | --- |
| **GO molecular functions** | | | |
| **Molecular function** | **Associated genes** | **P-value** | **FDR** |
| Complement component C3b binding (GO:0001851) | CFH, CFHR1, CFHR2, CFHR5 | 1.59 x 10^-09^ | 8.06 x 10^-06^ |
| Opsonin binding (GO:0001846) | CFH, CFHR1, CFHR2, CFHR5 | 1.47 x 10^-08^ | 3.71 x 10^-05^ |
| Complement binding (GO:0001848) | CFH, CFHR1, CFHR2, CFHR5 | 3.16 x 10^-08^ | 5.34 x 10^-05^ |
| **Reactome pathway** | | | |
| **Pathway** | **Associated genes** | **P-value** | **FDR** |
| Regulation of complement cascade | CFH, CFHR1, CFHR2, CFHR5 | 9.04 x 10^-06^ | 1.00 x 10^-03^ |
| Complement cascade | CFH, CFHR1, CFHR2, CFHR5 | 1.32 x 10^-05^ | 1.00 x 10^-03^ |
| **Variant rs6677604** | | | |
| **GO molecular functions** | | | |
| Complement component C3b binding (GO:0001851) | CFH, CFHR1 | 5.44 x 10^-06^ | 2.76 x 10^-02^ |
| **Reactome pathway** | | | |
| **Pathway** | **Associated genes** | **P-value** | **FDR** |
| Regulation of complement cascade | CFH, CFHR1 | 7.97 x 10^-05^ | 9.04 x 10^-04^ |
| Complement cascade | CFH, CFHR1 | 1.00 x 10^-04^ | 9.04 x 10^-04^ |
| MPS IIIB – Sanfilippo syndrome B | NAGLU | 6.76 x 10^-04^ | 0.004 |

CFH, complement factor H; CFHR, complement factor H related; FDR, false detection rate; GO, gene ontology; MPS IIIB, mucopolysaccharidosis type III B.
